# Digital seed amplification assay for TDP-43 aggregate quantification in CSF

**DOI:** 10.1101/2025.10.14.25338017

**Authors:** Ella Borberg, Zoe Swank, Tal Gilboa, Lily A. Wickland Shearer, Arek V. Melkonian, Justin C. Rolando, David R. Walt, Andrew M. Stern

**Author notes:** Correspondence: David R. Walt, Department of Pathology, Brigham and Women’s Hospital, Wyss Institute for Biologically Inspired Engineering, Harvard University, Harvard Medical School, Boston, MA, 02115, USA; Andrew M. Stern, Ann Romney Center for Neurologic Diseases, Department of Neurology, Brigham and Women’s Hospital, Harvard Medical School, Boston, Massachusetts, USA, 02115.

## Abstract

**INTRODUCTION:** Dementia is commonly caused by underlying pathologies driven by misfolded protein aggregates. Although dementia subtypes have distinct mechanisms, overlapping symptoms make diagnosis without biomarkers difficult. Misdiagnosis has previously hindered drug development by enrolling patients non-specifically in trials.

**METHODS:** We developed a digital Seed amplification assay (dSAA) that isolates individual aggregates in nanoliter compartments, enabling precise quantification of TDP-43 seeds in cerebrospinal fluid (CSF).

**RESULTS:** Testing 40 CSF samples from patients with genetic and sporadic FTLD-TDP, as well as healthy controls, we found elevated seed concentrations in FTLD-TDP patients that correlated with disease severity, demonstrating the potential of dSAA as a sensitive diagnostic tool.

**DISCUSSION:** This study demonstrates a new quantitative, high-sensitivity digital assay for TDP-43 seeds in CSF. The platform’s single-aggregate resolution and low LOD establish a technical foundation for developing a diagnostic and monitoring tool for FTLD-TDP and other TDP-43-related diseases.

## 1 BACKGROUND

As the global population ages, dementia, defined by progressive cognitive impairments that undermine daily activities, has become a growing social and economic issue, affecting over 20 million people worldwide, a number expected to double in the next 30 years.^1^ The most common dementia pathologies include Alzheimer’s disease (AD), Lewy body disease (LBD), Parkinson’s disease (PD), limbic-predominant age-related TDP-43 encephalopathy neuropathic change (LATE-NC), and frontotemporal lobar dementia with tau and TDP-43 subtypes (FTLD-tau and FTLD-TDP).^2,3^

These pathologies are defined by inclusions of aggregated proteins, such as amyloid-β, tau, α-synuclein, or TDP-43, that are signatures of neurodegeneration and cortical atrophy, presenting symptoms like behavioral changes, cognitive deficits, and motor dysfunction,^4–6^ several resulting in indistinguishable clinical phenotypes.^7,8^ Early AD clinical trials conducted without biomarker measurements enrolled a significant subset of patients that did not possess the underlying pathology of the drug target, contributing to drug development failure.^2,9^ Because only ∼45% of people with clinical behavioral variant frontotemporal dementia (bvFTD) exhibit intraneuronal pathological inclusions of TDP-43,^10^ untargeted enrollment of all bvFTD patients in a TDP-43-directed trial might result in failure of efficacious treatments for FTLD-TDP.^11^ Therefore, developing a test for FTLD-TDP has the potential to guide clinical trials and ultimately clinical practice.

Current diagnostic standards for FTLD-TDP include fluorine-18-fluorodeoxyglucose positron emission tomography (FDG-PET) and magnetic resonance imaging (MRI).^12^ Both generate useful information regarding regions of hypometabolism and of neurodegeneration, but they do not indicate the underlying biomarker pathology; therefore, they are unable to reliably distinguish between the different forms of FLTD.^13^ Efforts to measure total TDP-43 levels in plasma and CSF using immunoassays have not reproducibly succeeded at diagnosing FLTD-TDP.^14,15^ This difficulty may be due to the widespread physiologic abundance of TDP-43, which masks the relatively small fraction of its pathological aggregated form. Accurate detection requires assays that are sensitive to pathological TDP-43, capable of distinguishing the low concentrations released into CSF or plasma during the earliest stages of neurodegeneration.

In TDP-43 pathologies, cytoplasmic TDP-43 inclusions are thought to act as templates for the assembly of more fibrils.^16–18^ Seed amplification assays (SAAs) leverage this principle, using the patient’s fibrils as nucleation sites, or “seeds,” for aggregation, elongation, and amplification in the presence of recombinant monomers. When aggregates form, they bind to beta-sheet fluorophores,^19^ and generate a fluorescent signal.^20^ SAAs have the potential to be applied as early diagnostic screening tools for protein misfolding disorders and can provide valuable mechanistic insights into the aggregation of pathological protein variants.^21,22^ A leading example is α-synuclein SAAs used for diagnosis of synucleinopathy in CSF, with reported sensitivity of 84-95% and specificity of 68-95%.^23^

Using bulk SAA platforms, TDP-43 aggregates have been detected in cerebrospinal fluid (CSF); however, these assays lack quantitation capabilities,^24–26^ and spontaneous aggregation events of the monomer are amplified as false seeds, worsening the signal-to-noise ratios (SNRs).^27,28^ To facilitate both quantitation and avoid false positives, digital SAAs (dSAA) compartmentalize the bulk reaction into nanoliter volumes, the signals of these micro-compartments are counted in a binary on/off fashion, thereby digitizing the assay and producing a quantitative measure of “on” compartments, correlating with pathological seeds quantities in the sample.^29^ Therefore, dSAA enables single seed quantification, improving sensitivity, and limits spontaneous monomeric aggregation spread throughout the reaction.^30^

We previously developed a digital version of SAA for α-synuclein.^31^ Here, we developed a dSAA for quantifying pathological TDP-43 aggregates in CSF. We screened reaction conditions for optimal assay performance and implemented an image analysis pipeline to quantify the number of amplified aggregates. Then we next tested 40 CSF samples from 20 carriers with *GRN* or *C9ORF72* mutations that predict FTLD-TDP type A and B pathologies, respectively;^32–34^ 10 samples from non-carriers diagnosed with sporadic semantic variant of primary progressive aphasia (svPPA), predictive of FTLD-TDP type C pathology,^35^ and 10 samples of age-matched non-carrier healthy controls (CN). We found that FTLD-TDP patient CSF contained higher seed concentrations compared to CN participants, and seed concentrations increased with disease severity.

## 2 METHODS

### 2.1 Protein expression

N-terminal His-tagged expression plasmids were resuspended in TE buffer (Qiagen, #1018499) and transformed into Rosetta™ 2 (DE3) Competent Cells (Fisher Scientific, #713973) by heat shock. One-liter LB cultures were inoculated from single colonies and grown at 37 °C and shaking at 200 RPM to an OD_600_ of 0.6, then induced for target protein expression with 0.4 mM isopropyl β-D-1-thiogalactopyranoside at a final concentration overnight at 18 °C and shaking at 180 RPM. Cells were then harvested and centrifuged at 6,000 x *g,* then resuspended in binding buffer (20 mM Tris-HCl, 500 mM NaCl, 8 M urea, pH 8.0) with an additional 1% SDS, 1 mM 2-mercaptoethanol, and His-Tag Protease Inhibitor (Sigma #P8849) to assist with cell lysis. Nucleic acids were digested with >0.01 units/μL Benzonase, incubated for 30 minutes at 37 °C. Cells were further mechanically lysed through a 27G needle and centrifuged at 45,000 x *g* in a Beckman Type 70.1 rotor for 15 minutes at 4°C. The supernatant was diluted 1:1 in binding buffer and run through a His-trap Sepharose column (Cytiva, #17524802), and the protein was eluted via a linear imidazole gradient from 5 mM to 500 mM. The target protein was further purified using size exclusion chromatography (SEC) with an AKTA Pure™ protein purification system on a HiPrep 16/60 Sephacryl S-300 HR (Cytiva, #17116701) column with TBS (10 mM Tris, 500 mM NaCl), 4 M urea, pH 7.5 as the mobile phase. The relevant fractions were concentrated using a 10 kDa MWCO Amicon filter and stored at -80 °C.

### 2.2 Pre-formed fibril formation

Monomeric C-terminal fragment of human recombinant TDP-43 (TDP-C) was diluted to a concentration of 10 µM in TBS (20 mM Tris, 500 mM NaCl, pH 7.5) and shaken with 3mm Pyrex™ beads (Sigma-Aldrich, #CLS72683-1LB) at 600 rpm for 24-48 hours at 37 °C. Fibrils were then washed three times with subsequent 30-minute centrifugation steps at 185,000 x *g* in a Beckman TLA 55 rotor at 4°C; the pellet was resuspended each time in an equivalent volume of cold TBS. Samples were then stored at -80°C.

### 2.3 Transmission electron microscopy

TDP-C pre-formed fibrils samples were sonicated three times for 5 seconds, and 5 μL of 100 µM in monomer equivalent solution was incubated on top of glow-discharged carbon support film 400 mesh copper grids (Fisher Scientific, #50-248-91) for 60 seconds. Grids were then washed once with water and stained with 2% uranyl acetate for 30 seconds before removal of excess stain, followed by imaging on JOEL 1200EX using an AMT 2k CCD camera.

### 2.4 Clinical sample preparation

CSF was collected through the ARTFL LEFFTDS Longitudinal Frontotemporal Lobar Degeneration (ALLFTD) research network, following established protocols.^36^ In short, CSF was sampled with a spinal needle. Samples were transferred from collection tubes into polypropylene tubes, centrifuged, and frozen at -80 °C within 1 hour of sampling.

### 2.5 Microwell dSAA reaction

QuantStudio™ 3D Digital PCR 20K chips (Thermo Fisher Scientific) were prepared and used as previously described.^31^ SAA reaction components for the microwell arrays included 20 mM Tris (pH 6.5, Thermo Fisher Scientific), 100 mM Guanidine-Cl (Sigma-Aldrich), 10 mM DTT (Thermo Fisher Scientific), 0.0015% v/v SDS (Sigma-Aldrich), 1 µM TDP-C monomers, 0.1% v/v Triton-X 100 (Sigma-Aldrich), and 5 µM X-34 (Sigma-Aldrich). X-34 was dissolved in DMSO (Thermo Fisher Scientific) to a stock concentration of 6 mM, aliquoted, and stored at −20 °C. All prepared reagent solutions were filtered with 0.2 µm syringe filters before use (VWR). CSF samples were diluted tenfold into the SAA reaction.

### 2.6 Image acquisition

The chips were incubated on a heating block at 33 °C for 18 hours and were mounted onto a 3D-printed chip-holder for imaging. Images of the microwell array chips or droplets were acquired using a fluorescent microscope (Olympus IX83) equipped with an LED light source (CoolLED pE-300) and a 10× objective. The images were taken on each field of view with a reflected brightfield channel and the CFP fluorescence channel (Ex/Em 433/475 nm).

### 2.7 Image processing and data analysis

Performed using an automated custom Python (3.10) code and the scikit-image (0.19), NumPy (1.23), and pandas (1.4) packages. All image analysis code is available on GitHub (https://github.com/Wyss/chip-extract).^37^

### 2.8 Data interpolation and statistical analyses

GraphPad Prism 10.5.0 (774) was used to interpolate concentration values out of an on-well percentage calibration curve using a 4PL curve with 1/y^2^ weighting. All samples were blinded at the time of measurement and concentration interpolation. Measurements below LOD were assigned the LOD value. All statistical analyses were performed using GraphPad Prism 10.5.0 (774) software. In the pre-planned analysis, one-way ANOVA with post-doc Tukey’s test compared each FTLD-TDP group to the CN group. In post-hoc analyses, as follows, a lognormal unpaired two-tailed t-test was used to assess the significance of median variance between binned FTLD-TDP vs. control. Weighted multiple linear regression was used to model the relationship between seed concentration as an independent variable and sex at birth, age at CSF collection, genetic status, and FTLDCDR score as dependent variables. The choice to use weighing was made due to observed heteroskedasticity, **Figure S1**.^38,39^

## 3 RESULTS

### 3.1 Digital SAA optimization in buffer

SAA reactions combine monomeric peptides with pathological aggregates alongside a fluorescent dye that binds to the β-pleated sheet-rich amyloid fold. The pathological aggregates function as templates for misfolding, acting as seeds to attract the monomers,^40^ thereby amplifying the fluorescent signal, as shown schematically in **Figure 1A**. In our assays, the SAA reaction is compartmentalized into a microwell array containing 20,000 reaction wells, each comprising ∼0.8 nL volume. Monomers and the fluorescent dye are first added to a patient sample, and the reaction mixture is then compartmentalized into the microwell array and incubated at 33 °C. After 15 hours, the array is imaged to detect fluorescent wells corresponding to aggregate growth. Compartments are evaluated for fluorescent signals and determined to be either “on” or “off” (**Figure 1B**). We repurposed digital PCR microwell arrays fabricated from a silicon substrate (Applied Biosystems QuantStudio 3D Digital PCR 20K Chip).^41^ The chip’s layout is shown in **Figure 1C**. X-34 was used as the beta-sheet fluorophore. For initial assay development, the C-terminal fragment of human recombinant TDP-43, termed TDP-C, was used as the amplifying peptide, encompassing residues 252-414 with an N-terminal His-tag (**Figure 1D**).^42^ TDP-C lacks the TDP-43 N-terminal RNA-binding domains and contains a C-terminal fibril-forming low complexity domain, which is enriched in the fibrils found in human brain tissue.^43^ We first generated pre-formed fibrils (PFFs) from TDP-C to serve as reaction seeds; representative transmission electron microscopy images are shown in **Figure S2**. The PFF concentration presented is the concentration of monomers used to generate them (monomer equivalents). The exact molar quantity of seeds is difficult to estimate due to heterogeneous fibril lengths, but must be substantially lower than the monomer equivalent because each seed is comprised of more than one monomer.

**Figure 1.**
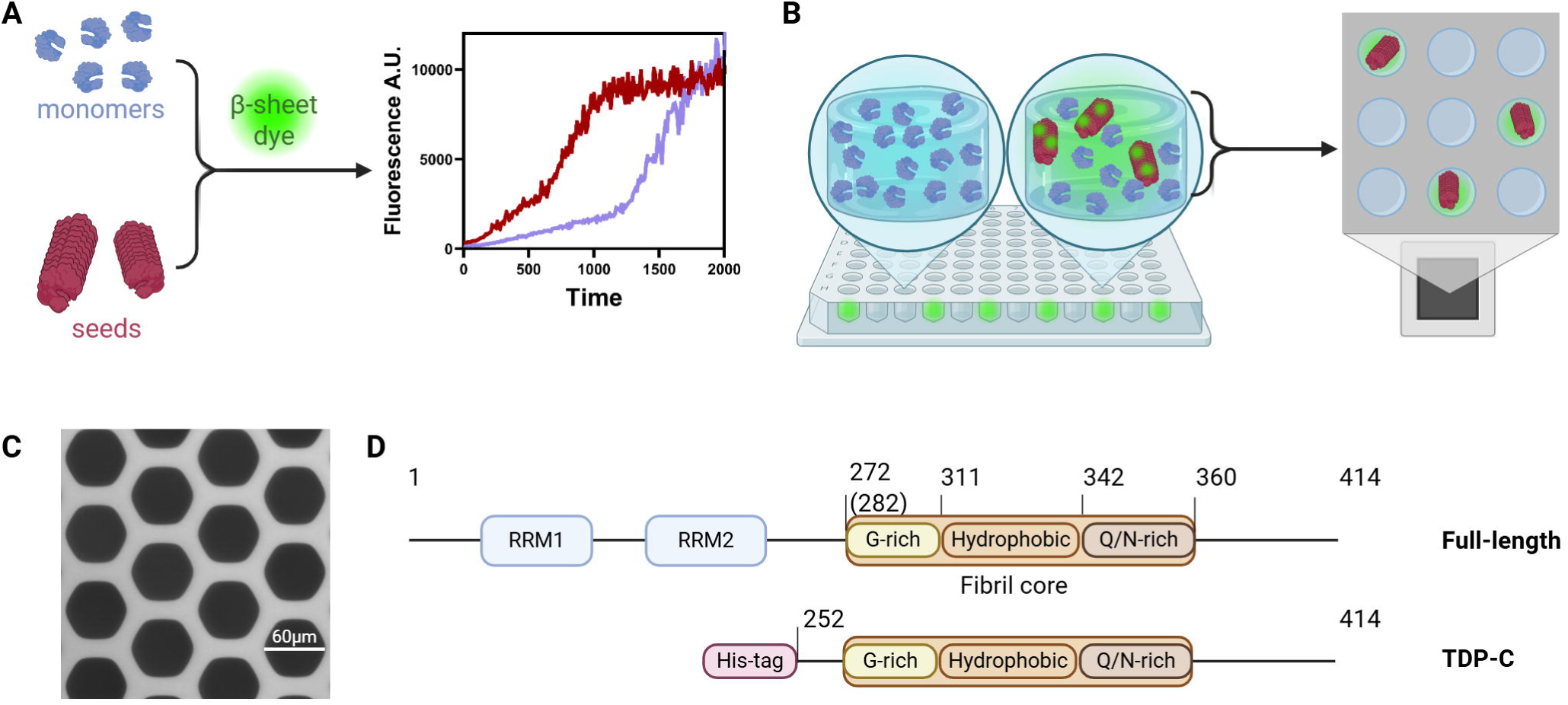
Digital seed amplification assay. **A.** Schematic of bulk SAA: pathological seeds are mixed with monomers and a beta-sheet fluorophore. **B.** For digitization, the same reaction components are partitioned into microcompartments of a microwell chip. **C.** Bright-field representative image of microwell array. **D.**TDP-43 full-length (top) and TDP-C peptide sequence (bottom). Created with biorender.com.

Reaction conditions for inducing TDP-43 aggregation, such as ionic strength, pH, and temperature, impact the rate and characteristics of the aggregation.^44,45^ We performed a set of experiments in which we explored various reaction parameters, including pH, buffers, surfactants, salts, polysaccharides, reducing agents, chemical denaturants, incubation temperature, and TDP-C monomer concentrations. As summarized in **Figure S3**, different conditions yield different SNRs, affecting both quantification and the limits of detection (LOD). The conditions most conducive to secondary nucleation were chosen for measuring biosamples: 1 μM TDP-C monomers, 0.1% v/v Triton-X, 10 mM dithiothreitol (DTT), 0.0015% v/v sodium dodecyl sulfate (SDS), 5 µM X-34, 20 mM Tris-Cl buffer, pH 6.5, incubated at 33 °C for 15 hours.

To generate calibration curves using the optimized reaction conditions, we spiked in PFFs ranging from 0.1-100 nM monomer equivalents into Tris-Cl buffer and quantified the percentage of wells containing growing aggregates in triplicate, **Figure 2**. Then, on-well percentages in each chip were determined using a custom Python image processing pipeline, **Figure S4**, analyzing approximately 13,500 wells per replicate. We determined that using PFFs spiked into Tris-Cl buffer, the monomer equivalent LOD and LLOQ are 41.0 pM and 1.25 nM, respectively.

**Figure 2.**
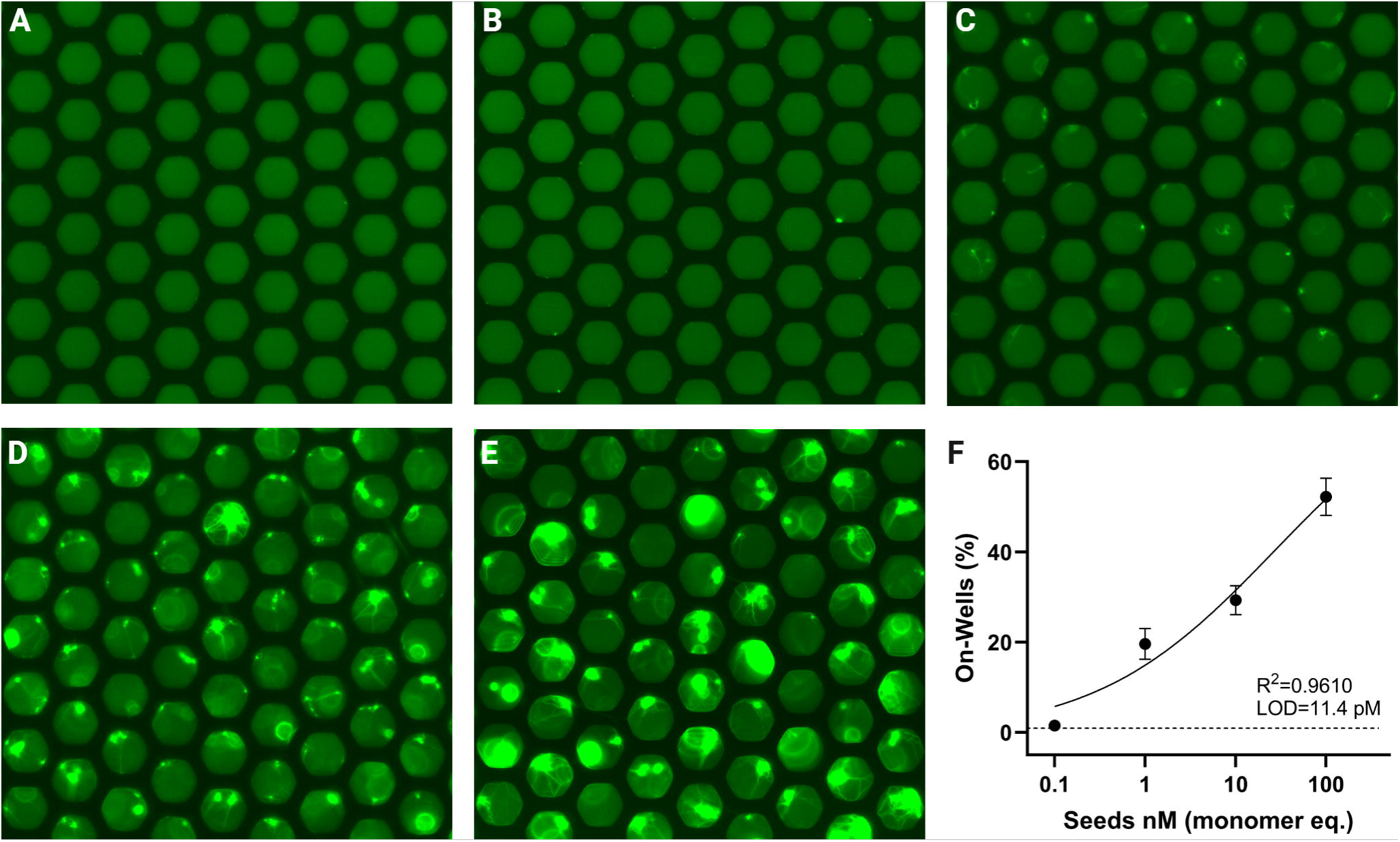
Synthetic TDP-C aggregate growth and quantification in buffer. Representative images of microwell arrays at t = 15 h. SAA reaction using PFFs as reaction seeds, PFFs concentrations of 0 nM **(A)**, 0.1 nM **(B)**, 1 nM **(C)**, 10 nM **(D)**, and 100 nM **(E)**. Note the increase in both number and size of aggregates with increasing starting monomer concentration. The percentage of wells with growing aggregates vs. the PFFs concentrations used as reaction seeds spiked into Tris-Cl buffer, trend line shows the 4PL curve **(F)**.

### 3.2 Digital SAA optimization in biosamples

Our previous work^31^ has shown that reaction conditions affect SNR and adjustments for different biomatrices are required. After achieving sufficient specificity and sensitivity in buffer conditions using PFFs, we further optimized the dSAA using brain lysates and CSF derived from healthy individuals. We tested various reaction parameters, including protein concentration, incubation time, and reagent composition, to counteract matrix effects. The resulting calibration curve in CSF is shown in **Figure 3**, and the resulting calibration curve in brain lysate is shown in **Figure S5**. From these curves, we determined that using PFFs spiked into brain lysate, the monomer equivalent LOD and LLOQ are 172 pM and 3.66 nM, respectively, and using PFFs spiked into CSF, the LOD and LLOQ are 1.32 nM and 6.41 nM, respectively.

**Figure 3.**
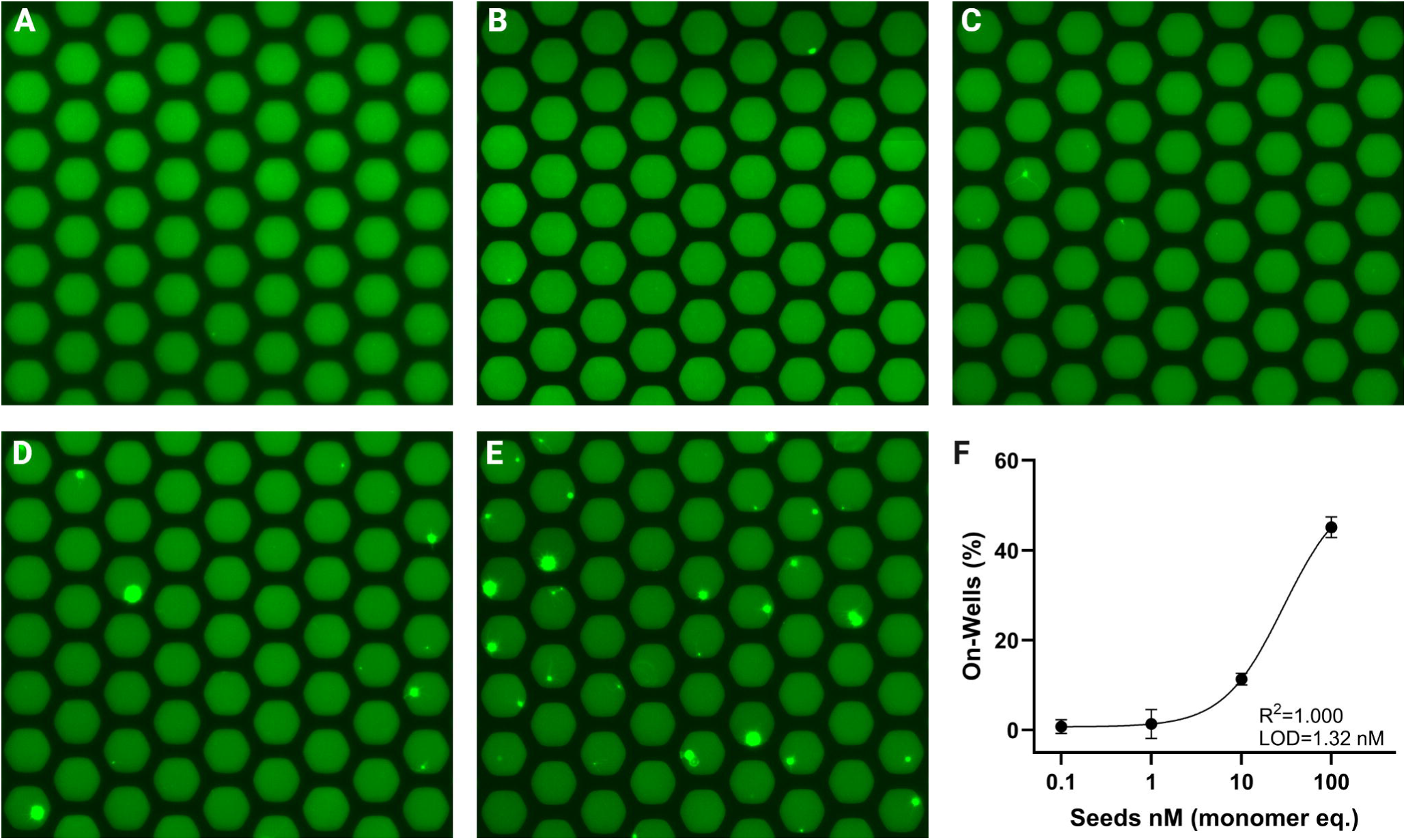
Synthetic TDP-C aggregate growth and quantification in CSF. Representative images of microwell arrays at t = 15 h. SAA reaction using PFFs as reaction seeds, PFFs concentrations of 0 nM **(A)**, 0.1 nM **(B)**, 1 nM **(C)**, 10 nM **(D)**, and 100 nM **(E)**. The percentage of wells with growing aggregates vs. the PFFs concentrations used as reaction seeds spiked into CSF, trend line shows the 4PL curve **(F)**.

### 3.3 Quantification of pathological TDP-43 aggregates in patient samples

After optimizing the reaction conditions, we tested 40 CSF samples from the ALLFTD cohort, representing three groups of different FTLD-TDP subtypes: *GRN* mutation carriers, *C9ORF72* repeat expansion carriers, patients without mutations but with a clinical diagnosis of svPPA, and a control group (CN). Patient characteristics are summarized in **Table 1**. This design enabled us to assess the performance of the assay across FTLD-TDP pathological subtypes. In total, 30 out of 40 samples were successfully detected above the assay’s LOD. Mean seed concentrations of triplicate chips per sample were grouped by FTLD-TDP subtype in our pre-planned analysis. We found that *GRN* carriers possessed higher seed concentrations compared to NC; we did not detect significantly higher seed concentrations in *C9ORF72* or svPPA CSF compared to CN, although the group’s median seed concentrations were numerically higher, **Figure 4A**. The specific mean concentrations, replicate measurement values, and their corresponding coefficient of variation percentages are reported in **Table S1**.

**Figure 4.**
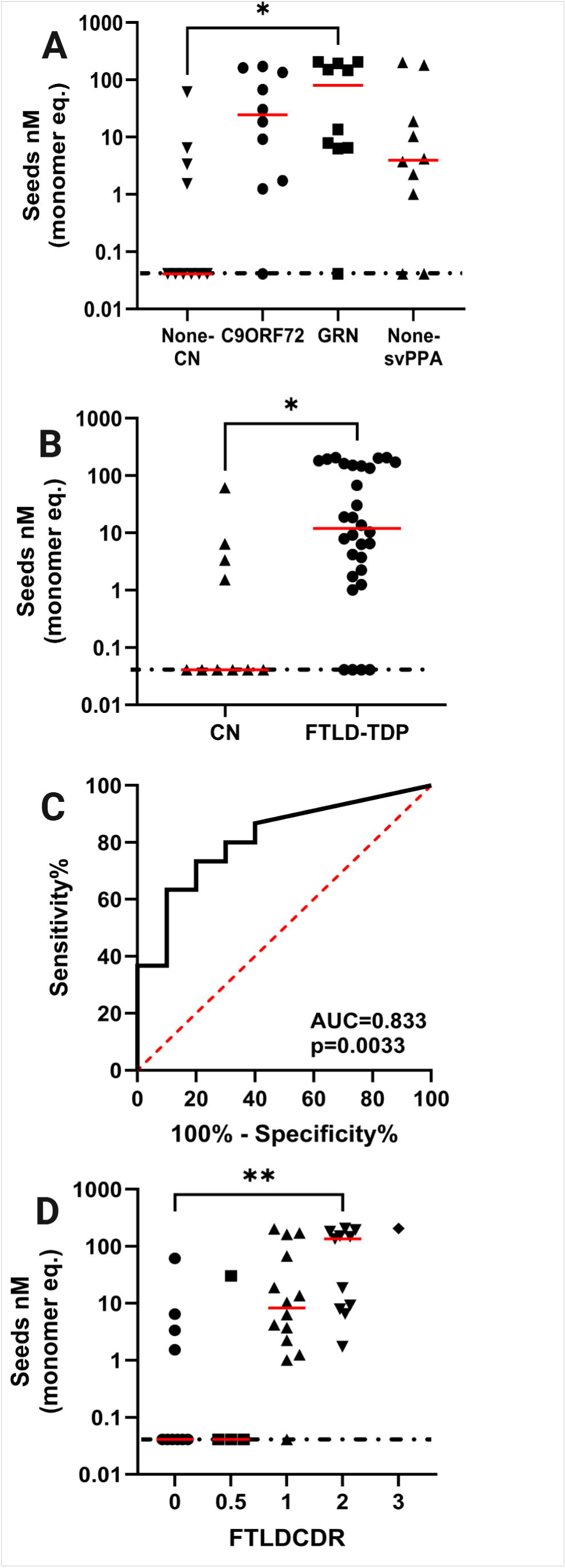
Seed concentrations vs. FLTD status and severity. **A.** Observed seed concentrations of samples grouped by FLTD subtypes. **B.** Observed seed concentrations of FLTD-positive patients and CN participants. **C.** ROC curve presenting the diagnostic performance of seed concentration for distinguishing FTLD vs control. **D.** Observed seed concentrations of samples grouped by FTLDCDR severity scores. Red horizontal lines represent the group median value. Dotted horizontal line represents assay LOD. To assess the significance of median variance between binned categories vs. controls, a Lognormal t-test was used for **B,** and one-way ANOVA with post-doc Tukey’s test was used for **A** and **D.**

**Table 1.** Cohort characteristics.

In a post-hoc analysis, when combining all three FTLD-TDP groups into one, we found an overall significantly higher concentration of seeds in FTLD-TDP samples compared to controls (**Figure 4B**, *p =* 0.0334). Using receiver operating characteristic (ROC) curve analysis, **Figure 4C**, the optimal cutoff value to differentiate FLTD-TDP from non-TDP pathologies was determined to be a seed monomer equivalent of 0.528 nM. With this cutoff, we calculated a sensitivity of 87%, specificity of 60%, positive predictive value of 87%, and negative predictive value of 60%, **Table 2**.

**Table 2.** Assay performance.

### 3.4 Statistical examination of TDP-43 aggregates in patient samples

To assess the correlation between disease severity and seed concentrations, we correlated the CDR plus NACC (clinical dementia rating plus National Alzheimer’s Coordination Center) FTLD scores, denoted FTLDCDR, which have been reported to track the clinical progression of FLTD.^46–48^ The FTLDCDR is rated on a five-point ordinal scale starting from clinically normal (0), minimally impaired (0.5), mildly impaired (1), moderately impaired (2), to severely impaired (3). Median seed concentrations correlated with FTLDCDR, **Figure 4D**. Simple linear regression, **Figure S6**, showed a significant relationship between seed concentration and FLTDCDR (*p* = 0.0002, R^2^ = 0.308). Although there is a correlation between seed quantification and median FTLDCDR scores, the wide range of values for individual patients suggests it will be necessary to measure seed concentrations at multiple time points if the assay is to be used to follow disease progression. When comparing seed concentrations of primary clinical phenotype, the median was not significantly different, **Figure S7**.

We also conducted a multiple linear regression (MLR) analysis to evaluate the relationship of seed concentration with pathologic diagnosis and FTLDCDR, accounting for age and sex. This analysis demonstrated a multiple R of 0.620 and a weighted R^2^ of 0.384. MLR performance parameters are listed in **Table 3**. The overall model was statistically significant (F(6, 33) = 3.43, *p* = 0.0097). Analysis of variance, including F-statistics and associated *p*-values, is shown in **Table 4**. Additional performance assessment plots of the MLR are presented in **Figure S8** and a summary variance inflation factor is in **Table S2**. Sex, FTLD-TDP subtype, and FTLDCDR were all significantly associated with seed concentration. Female sex was associated with a lower seed concentration, and higher FTLDCDR scores were associated with higher seed concentration. Age at CSF collection was not a significant predictor.

**Table 3.** MLR model performance, using seed concentration as the dependent variable.

**Table 4.** MLR variable performance, using seed concentration as the dependent variable.

## 4 DISCUSSION

Establishing a sensitive, quantitative, and accurate test for TDP-43 aggregates would be useful as a biomarker assay to screen patients, to monitor treatment efficacy during clinical trials, and to follow disease progression. In this work, we developed a digital SAA for quantifying TDP-43 aggregates in CSF. To digitize the SAA readout, we confined diluted CSF into microwells, enabling us to quantify the number of pathological aggregates in the sample. By combining a highly sensitive digital assay with programmed image analysis, we aimed to overcome limitations of traditional bulk assays, such as false positives and a lack of quantitation. We found our dSAA to be a sensitive quantification platform for TDP-43 aggregates, with an LOD = 41 pM in monomer equivalent.

After finding optimal assay conditions, we applied the dSAA to a preliminary cohort of 40 CSF samples, including 10 carriers of *C9ORF72* repeat expansions, 10 carriers of *GRN* mutations (both strong genetic predictors of FTLD-TDP pathology), 10 patients with sporadic svPPA (a predictor of FTLD-TDP pathology), and 10 age-matched clinically normal non-carriers (CN).^49–52^ In our pre-planned analysis dividing the FTLD-TDP groups, we found that *GRN* carriers had significantly higher seed concentrations compared to CN controls. In a post-hoc analysis, we found that the FTLD-TDP group as a whole had significantly higher seed concentrations compared to CN, and the concentrations increased significantly with disease severity as measured by FTLDCDR scores.

While these results are promising, this study has several limitations. First, the small sample size limits the statistical power. Second, conducting a ROC analysis to establish sensitivities and specificities is limited because of the lack of a validation cohort. Third, most patients in our cohort have unconfirmed pathological classifications of FLTD-TDP by autopsy; we note that one of the two false-positive CN participants who exhibited higher seed concentration had a single abnormal neurological evaluation exam at one time point. Fourth, the cross-sectional nature meant that we could not determine the effect of within-subject disease progression on CSF seed concentration. Fifth, we did not evaluate a diseased, non-FTLD-TDP group (such as an FTLD-tau group), hindering evaluation for cross-seeding due to tau aggregates in CSF. Sixth, our calibrator was derived from *in vitro* PFFs without the ability to determine their molar concentration, making a precise estimate of the molar concentration of seeds in patient samples challenging. Seventh, the adaptation of digital PCR chips and a custom image analysis pipeline makes the assay more difficult to reproduce than a typical immunoassay on a commercial platform, but our analysis code is shared publicly.

Despite these limitations, this TDP-43 dSAA shows promise. It should be evaluated in larger, pathologically confirmed FTLD cohorts, with separate discovery and validation groups, as well as cohorts with confirmed ALS, LATE-NC, and FTLD-tau pathology. Longitudinal sampling will help validate the assay’s ability to detect changes over time and determine its value for staging disease or monitoring therapeutic response. Overall, this study demonstrates a new quantitative, high-sensitivity digital assay for TDP-43 seeds in CSF. The platform’s single-aggregate resolution and low LOD establish a technical foundation for developing a diagnostic and monitoring tool for FTLD-TDP and other TDP-43-related diseases.

## Data Availability

All data produced in the present study are available upon reasonable request to the authors.

## ACKNOWLEDGMENTS

We thank Yoshiaki Furukawa for providing plasmids encoding TDP-C.

Data collection and dissemination of the data presented in this manuscript was supported by the ALLFTD Consortium (U19: AG063911, funded by the National Institute on Aging and the National Institute of Neurological Diseases and Stroke) and the former ARTFL & LEFFTDS Consortia (ARTFL: U54 NS092089, funded by the National Institute of Neurological Diseases and Stroke and National Center for Advancing Translational Sciences; LEFFTDS: U01 AG045390, funded by the National Institute on Aging and the National Institute of Neurological Diseases and Stroke). The authors acknowledge the invaluable contributions of the study participants and families, as well as the assistance of the support staff at each of the participating sites.

## AUTHOR CONTRIBUTIONS

All authors contributed to the study conception and design. Material preparation was performed by Ella Borberg, Lily A. Wickland Shearer, and Arek V. Melkonian. Data collection was performed by Ella Borberg, Zoe Swank, Tal Gilboa, and Lily A. Wickland Shearer. Data analysis was performed by Ella Borberg, Tal Gilboa, and Justin C. Rolando. The first draft of the manuscript was written by Ella Borberg, and all authors commented on previous versions of the manuscript. All authors read and approved the final manuscript.

## DATA AVAILABILITY

The data supporting the findings of this study are available within the paper and its Supplementary Information. Should any raw data files be needed, they would be shared upon reasonable request from the corresponding author. Python codes have been deposited to Github (https://github.com/Wyss/chip-extract).^37^

## CONSENT STATEMENT

All participants and/or their legally authorized representatives provided written informed consent under the National Centralized Repository for Alzheimer’s Disease and Related Dementias (NCRAD) Biospecimen Review Committee.

## REFERENCES

(1) Heutink, P. Untangling Tau-Related Dementia. Human Molecular Genetics 2000, 9 (6), 979–986. 10.1093/hmg/9.6.979.

(2) Schneider, L. S.; Mangialasche, F.; Andreasen, N.; Feldman, H.; Giacobini, E.; Jones, R.; Mantua, V.; Mecocci, P.; Pani, L.; Winblad, B.; Kivipelto, M. Clinical Trials and Late-Stage Drug Development for Alzheimer’s Disease: An Appraisal from 1984 to 2014. Journal of Internal Medicine 2014, 275 (3), 251–283. 10.1111/joim.12191.

(3) Walker, L. C.; Jucker, M. Neurodegenerative Diseases: Expanding the Prion Concept. Annual Review of Neuroscience 2015, 38 (Volume 38, 2015), 87–103. 10.1146/annurev-neuro-071714-033828.

(4) Breijyeh, Z.; Karaman, R. Comprehensive Review on Alzheimer’s Disease: Causes and Treatment. Molecules 2020, 25 (24), 5789. 10.3390/molecules25245789.

(5) Ghetti, B.; Oblak, A. L.; Boeve, B. F.; Johnson, K. A.; Dickerson, B. C.; Goedert, M. Invited Review: Frontotemporal Dementia Caused by Microtubule-Associated Protein Tau Gene (MAPT) Mutations: A Chameleon for Neuropathology and Neuroimaging. Neuropathol Appl Neurobiol 2015, 41 (1), 24–46. 10.1111/nan.12213.

(6) Grossman, M.; Seeley, W. W.; Boxer, A. L.; Hillis, A. E.; Knopman, D. S.; Ljubenov, P. A.; Miller, B.; Piguet, O.; Rademakers, R.; Whitwell, J. L.; Zetterberg, H.; van Swieten, J. C. Frontotemporal Lobar Degeneration. Nat Rev Dis Primers 2023, 9 (1), 1–19. 10.1038/s41572-023-00447-0.

(7) Jellinger, K. A. Recent Update on the Heterogeneity of the Alzheimer’s Disease Spectrum. J Neural Transm 2022, 129 (1), 1–24. 10.1007/s00702-021-02449-2.

(8) Moda, F.; Ciullini, A.; Dellarole, I. L.; Lombardo, A.; Campanella, N.; Bufano, G.; Cazzaniga, F. A.; Giaccone, G. Secondary Protein Aggregates in Neurodegenerative Diseases: Almost the Rule Rather than the Exception. FBL 2023, 28 (10), 255. 10.31083/j.fbl2810255.

(9) Kim, C. K.; Lee, Y. R.; Ong, L.; Gold, M.; Kalali, A.; Sarkar, J. Alzheimer’s Disease: Key Insights from Two Decades of Clinical Trial Failures. Journal of Alzheimer’s Disease 2022, 87 (1), 83–100. 10.3233/JAD-215699.

(10) Frontotemporal degeneration, the next therapeutic frontier: Molecules and animal models for frontotemporal degeneration drug development - Boxer - 2013 - Alzheimer’s & Dementia - Wiley Online Library. https://alz-journals.onlinelibrary.wiley.com/doi/10.1016/j.jalz.2012.03.002 (accessed 2025-04-30).

(11) Panza, F.; Lozupone, M.; Seripa, D.; Daniele, A.; Watling, M.; Giannelli, G.; Imbimbo, B. P. Development of Disease-Modifying Drugs for Frontotemporal Dementia Spectrum Disorders. Nat Rev Neurol 2020, 16 (4), 213–228. 10.1038/s41582-020-0330-x.

(12) Rascovsky, K.; Hodges, J. R.; Knopman, D.; Mendez, M. F.; Kramer, J. H.; Neuhaus, J.; van Swieten, J. C.; Seelaar, H.; Dopper, E. G. P.; Onyike, C. U.; Hillis, A. E.; Josephs, K. A.; Boeve, B. F.; Kertesz, A.; Seeley, W. W.; Rankin, K. P.; Johnson, J. K.; Gorno-Tempini, M.-L.; Rosen, H.; Prioleau-Latham, C. E.; Lee, A.; Kipps, C. M.; Lillo, P.; Piguet, O.; Rohrer, J. D.; Rossor, M. N.; Warren, J. D.; Fox, N. C.; Galasko, D.; Salmon, D. P.; Black, S. E.; Mesulam, M.; Weintraub, S.; Dickerson, B. C.; Diehl-Schmid, J.; Pasquier, F.; Deramecourt, V.; Lebert, F.; Pijnenburg, Y.; Chow, T. W.; Manes, F.; Grafman, J.; Cappa, S. F.; Freedman, M.; Grossman, M.; Miller, B. L. Sensitivity of Revised Diagnostic Criteria for the Behavioural Variant of Frontotemporal Dementia. Brain 2011, 134 (9), 2456–2477. 10.1093/brain/awr179.

(13) López-Carbonero, J. I.; García-Toledo, I.; Fernández-Hernández, L.; Bascuñana, P.; Gil-Moreno, M. J.; Matías-Guiu, J. A.; Corrochano, S. In Vivo Diagnosis of TDP-43 Proteinopathies: In Search of Biomarkers of Clinical Use. Transl Neurodegener 2024, 13 (1), 29. 10.1186/s40035-024-00419-8.

(14) Chatterjee, M.; Özdemir, S.; Fritz, C.; Möbius, W.; Kleineidam, L.; Mandelkow, E.; Biernat, J.; Doğdu, C.; Peters, O.; Cosma, N. C.; Wang, X.; Schneider, L.-S.; Priller, J.; Spruth, E.; Kühn, A. A.; Krause, P.; Klockgether, T.; Vogt, I. R.; Kimmich, O.; Spottke, A.; Hoffmann, D. C.; Fliessbach, K.; Miklitz, C.; McCormick, C.; Weydt, P.; Falkenburger, B.; Brandt, M.; Guenther, R.; Dinter, E.; Wiltfang, J.; Hansen, N.; Bähr, M.; Zerr, I.; Flöel, A.; Nestor, P. J.; Düzel, E.; Glanz, W.; Incesoy, E.; Bürger, K.; Janowitz, D.; Perneczky, R.; Rauchmann, B. S.; Hopfner, F.; Wagemann, O.; Levin, J.; Teipel, S.; Kilimann, I.; Goerss, D.; Prudlo, J.; Gasser, T.; Brockmann, K.; Mengel, D.; Zimmermann, M.; Synofzik, M.; Wilke, C.; Selma-González, J.; Turon-Sans, J.; Santos-Santos, M. A.; Alcolea, D.; Rubio-Guerra, S.; Fortea, J.; Carbayo, Á.; Lleó, A.; Rojas-García, R.; Illán-Gala, I.; Wagner, M.; Frommann, I.; Roeske, S.; Bertram, L.; Heneka, M. T.; Brosseron, F.; Ramirez, A.; Schmid, M.; Beschorner, R.; Halle, A.; Herms, J.; Neumann, M.; Barthélemy, N. R.; Bateman, R. J.; Rizzu, P.; Heutink, P.; Dols-Icardo, O.; Höglinger, G.; Hermann, A.; Schneider, A. Plasma Extracellular Vesicle Tau and TDP-43 as Diagnostic Biomarkers in FTD and ALS. Nat Med 2024, 30 (6), 1771–1783. 10.1038/s41591-024-02937-4.

(15) Feneberg, E.; Gray, E.; Ansorge, O.; Talbot, K.; Turner, M. R. Towards a TDP-43-Based Biomarker for ALS and FTLD. Mol Neurobiol 2018, 55 (10), 7789–7801. 10.1007/s12035-018-0947-6.

(16) Gibbons, G. S.; Lee, V. M. Y.; Trojanowski, J. Q. Mechanisms of Cell-to-Cell Transmission of Pathological Tau: A Review. JAMA Neurology 2019, 76 (1), 101–108. 10.1001/jamaneurol.2018.2505.

(17) Goedert, M.; Eisenberg, D. S.; Crowther, R. A. Propagation of Tau Aggregates and Neurodegeneration. Annual Review of Neuroscience 2017, 40 (Volume 40, 2017), 189–210. 10.1146/annurev-neuro-072116-031153.

(18) Versluys, L.; Ervilha Pereira, P.; Schuermans, N.; De Paepe, B.; De Bleecker, J. L.; Bogaert, E.; Dermaut, B. Expanding the TDP-43 Proteinopathy Pathway From Neurons to Muscle: Physiological and Pathophysiological Functions. Front. Neurosci. 2022, 16. 10.3389/fnins.2022.815765.

(19) Styren, Scot D.; Hamilton, R. L.; Styren, G. C.; Klunk, W. E. X-34, A Fluorescent Derivative of Congo Red: A Novel Histochemical Stain for Alzheimer’s Disease Pathology. J Histochem Cytochem. 2000, 48 (9), 1223–1232. 10.1177/002215540004800906.

(20) Srivastava, A.; Singh, P. K.; Kumbhakar, M.; Mukherjee, T.; Chattopadyay, S.; Pal, H.; Nath, S. Identifying the Bond Responsible for the Fluorescence Modulation in an Amyloid Fibril Sensor. Chemistry A European J 2010, 16 (30), 9257–9263. 10.1002/chem.200902968.

(21) Rossi, M.; Candelise, N.; Baiardi, S.; Capellari, S.; Giannini, G.; Orrù, C. D.; Antelmi, E.; Mammana, A.; Hughson, A. G.; Calandra-Buonaura, G.; Ladogana, A.; Plazzi, G.; Cortelli, P.; Caughey, B.; Parchi, P. Correction to: Ultrasensitive RT-QuIC Assay with High Sensitivity and Specificity for Lewy Body-Associated Synucleinopathies. Acta Neuropathol 2020, 140 (2), 245–245. 10.1007/s00401-020-02170-6.

(22) Rossi, M.; Candelise, N.; Baiardi, S.; Capellari, S.; Giannini, G.; Orrù, C. D.; Antelmi, E.; Mammana, A.; Hughson, A. G.; Calandra-Buonaura, G.; Ladogana, A.; Plazzi, G.; Cortelli, P.; Caughey, B.; Parchi, P. Ultrasensitive RT-QuIC Assay with High Sensitivity and Specificity for Lewy Body-Associated Synucleinopathies. Acta Neuropathol 2020, 140 (1), 49–62. 10.1007/s00401-020-02160-8.

(23) Ma, Y.; Farris, C. M.; Weber, S.; Schade, S.; Nguyen, H.; Pérez-Soriano, A.; Giraldo, D. M.; Fernández, M.; Soto, M.; Cámara, A.; Painous, C.; Muñoz, E.; Valldeoriola, F.; Martí, M. J.; Clarimon, J.; Kallunki, P.; Ma, T. C.; Alcalay, R. N.; Gomes, B. F.; Blennow, K.; Zetterberg, H.; Constantinescu, J.; Mengel, D.; Kadam, V.; Parchi, P.; Brockmann, K.; Tropea, T. F.; Siderowf, A.; Synofzik, M.; Kang, U. J.; Compta, Y.; Svenningsson, P.; Mollenhauer, B.; Concha-Marambio, L. Sensitivity and Specificity of a Seed Amplification Assay for Diagnosis of Multiple System Atrophy: A Multicentre Cohort Study. The Lancet Neurology 2024, 23 (12), 1225–1237. 10.1016/s1474-4422(24)00395-8.

(24) Audrain, M.; Egesipe, A.-L.; Tentillier, N.; Font, L.; Ratnam, M.; Mottier, L.; Clavel, M.; Le Roux-Bourdieu, M.; Fenyi, A.; Ollier, R.; Chevalier, E.; Guilhot, F.; Fuchs, A.; Piorkowska, K.; Carlyle, B.; Arnold, S. E.; Berry, J. D.; Luthi-Carter, R.; Adolfsson, O.; Pfeifer, A.; Kosco-Vilbois, M.; Seredenina, T.; Afroz, T. Targeting Amyotrophic Lateral Sclerosis by Neutralizing Seeding-Competent TDP-43 in CSF. Brain Communications 2023, 5 (6), fcad306. 10.1093/braincomms/fcad306.

(25) Scialò, C.; Tran, T. H.; Salzano, G.; Novi, G.; Caponnetto, C.; Chiò, A.; Calvo, A.; Canosa, A.; Moda, F.; Caroppo, P.; Silani, V.; Ticozzi, N.; Ratti, A.; Borroni, B.; Benussi, L.; Ghidoni, R.; Furlanis, G.; Manganotti, P.; Senigagliesi, B.; Parisse, P.; Brasselet, R.; Buratti, E.; Legname, G. TDP-43 Real-Time Quaking Induced Conversion Reaction Optimization and Detection of Seeding Activity in CSF of Amyotrophic Lateral Sclerosis and Frontotemporal Dementia Patients. Brain Commun 2020, 2 (2), fcaa142. 10.1093/braincomms/fcaa142.

(26) Fontana, E.; Bongianni, M.; Benussi, A.; Bronzato, E.; Scialo, C.; Sacchetto, L.; Cagnin, A.; Castriciano, S.; Buratti, E.; Gardoni, F.; Italia, M.; Schreiber, A.; Ferracin, C.; Fiorini, M.; Newell, K. L.; Cracco, L.; Garringer, H. J.; Cecchini, M. P.; Polymenidou, M.; Padovani, A.; Monaco, S.; Legname, G.; Ghetti, B.; Borroni, B.; Zanusso, G. Detection of TDP-43 Seeding Activity in the Olfactory Mucosa from Patients with Frontotemporal Dementia. Alzheimer’s & Dementia 2024, 20 (2), 1156–1165. 10.1002/alz.13541.

(27) Cordts, I.; Wachinger, A.; Scialo, C.; Lingor, P.; Polymenidou, M.; Buratti, E.; Feneberg, E. TDP-43 Proteinopathy Specific Biomarker Development. Cells 2023, 12 (4), 597. 10.3390/cells12040597.

(28) Paciotti, S.; Bellomo, G.; Gatticchi, L.; Parnetti, L. Are We Ready for Detecting α-Synuclein Prone to Aggregation in Patients? The Case of “Protein-Misfolding Cyclic Amplification” and “Real-Time Quaking-Induced Conversion” as Diagnostic Tools. Front. Neurol. 2018, 9, 415. 10.3389/fneur.2018.00415.

(29) Pfammatter, M.; Andreasen, M.; Meisl, G.; Taylor, C. G.; Adamcik, J.; Bolisetty, S.; Sánchez-Ferrer, A.; Klenerman, D.; Dobson, C. M.; Mezzenga, R.; Knowles, T. P. J.; Aguzzi, A.; Hornemann, S. Absolute Quantification of Amyloid Propagons by Digital Microfluidics. Anal. Chem. 2017, 89 (22), 12306–12313. 10.1021/acs.analchem.7b03279.

(30) Coysh, T.; Mead, S. The Future of Seed Amplification Assays and Clinical Trials. Front. Aging Neurosci. 2022, 14. 10.3389/fnagi.2022.872629.

(31) Gilboa, T.; Swank, Z.; Thakur, R.; Gould, R. A.; Ooi, K. H.; Norman, M.; Flynn, E. A.; Deveney, B. T.; Chen, A.; Borberg, E.; Kuzkina, A.; Ndayisaba, A.; Khurana, V.; Weitz, D. A.; Walt, D. R. Toward the Quantification of α-Synuclein Aggregates with Digital Seed Amplification Assays. Proc. Natl. Acad. Sci. U.S.A. 2024, 121 (3), e2312031121. 10.1073/pnas.2312031121.

(32) Bennion Callister, J.; Pickering-Brown, S. M. Pathogenesis/Genetics of Frontotemporal Dementia and How It Relates to ALS. Experimental Neurology 2014, 262, 84–90. 10.1016/j.expneurol.2014.06.001.

(33) Gijselinck, I.; Van Broeckhoven, C.; Cruts, M. Granulin Mutations Associated with Frontotemporal Lobar Degeneration and Related Disorders: An Update. Human Mutation 2008, 29 (12), 1373–1386. 10.1002/humu.20785.

(34) Pottier, C.; Mateiu, L.; Baker, M. C.; DeJesus-Hernandez, M.; Teixeira Vicente, C.; Finch, N. A.; Tian, S.; van Blitterswijk, M.; Murray, M. E.; Ren, Y.; Petrucelli, L.; Oskarsson, B.; Biernacka, J. M.; Graff-Radford, N. R.; Boeve, B. F.; Petersen, R. C.; Josephs, K. A.; Asmann, Y. W.; Dickson, D. W.; Rademakers, R. Shared Brain Transcriptomic Signature in TDP-43 Type A FTLD Patients with or without GRN Mutations. Brain 2022, 145 (7), 2472– 2485. 10.1093/brain/awab437.

(35) Mackenzie, I. R. A.; Neumann, M.; Baborie, A.; Sampathu, D. M.; Plessis, D. D.; Jaros, E.; Perry, R. H.; Trojanowski, J. Q.; Mann, D. M. A.; Lee, V. M. Y. A Harmonized Classification System for FTLD-TDP Pathology. Acta Neuropathol 2011, 122 (1), 111–113. 10.1007/s00401-011-0845-8.

(36) Edler, M. C.; Faber, K.; Lacy, K.; Jackson, J.; Foroud, T. NCRAD: Advancing Alzheimer’s Research through High-Quality Biospecimens and Data. Alzheimer’s & Dementia 2025, 21 (9), e70682. 10.1002/alz.70682.

(37) Wyss/Chip-Extract, 2024. https://github.com/Wyss/chip-extract (accessed 2025-08-26).

(38) Bolic, M. Chapter 1 - Concepts in Performance Evaluation and Uncertainty Analysis. In Pervasive Cardiovascular and Respiratory Monitoring Devices; Bolic, M., Ed.; Academic Press, 2023; pp 1–35. 10.1016/B978-0-12-820947-9.00008-8.

(39) Foroud, T.; Faber, K.; Kim, C.; Dickinson, J. ALLFTD Manual of Procedures Update. https://ncrad.iu.edu/assets/docs/resource/allftd/Manual_of_Procedures_ALLFTD_NCRAD_04_2025.pdf (accessed 2025-01-08).

(40) Louros, N.; Schymkowitz, J.; Rousseau, F. Mechanisms and Pathology of Protein Misfolding and Aggregation. Nat Rev Mol Cell Biol 2023, 24 (12), 912–933. 10.1038/s41580-023-00647-2.

(41) Rolando, J. C.; Jue, E.; Schoepp, N. G.; Ismagilov, R. F. Real-Time, Digital LAMP with Commercial Microfluidic Chips Reveals the Interplay of Efficiency, Speed, and Background Amplification as a Function of Reaction Temperature and Time. Anal. Chem. 2019, 91 (1), 1034–1042. 10.1021/acs.analchem.8b04324.

(42) Furukawa, Y.; Kaneko, K.; Watanabe, S.; Yamanaka, K.; Nukina, N. A Seeding Reaction Recapitulates Intracellular Formation of Sarkosyl-Insoluble Transactivation Response Element (TAR) DNA-Binding Protein-43 Inclusions. J Biol Chem 2011, 286 (21), 18664–18672. 10.1074/jbc.M111.231209.

(43) Kametani, F.; Obi, T.; Shishido, T.; Akatsu, H.; Murayama, S.; Saito, Y.; Yoshida, M.; Hasegawa, M. Mass Spectrometric Analysis of Accumulated TDP-43 in Amyotrophic Lateral Sclerosis Brains. Sci Rep 2016, 6, 23281. 10.1038/srep23281.

(44) Jeganathan, S.; von Bergen, M.; Mandelkow, E.-M.; Mandelkow, E. The Natively Unfolded Character of Tau and Its Aggregation to Alzheimer-like Paired Helical Filaments. Biochemistry 2008, 47 (40), 10526–10539. 10.1021/bi800783d.

(45) Santos, J.; Iglesias, V.; Santos-Suárez, J.; Mangiagalli, M.; Brocca, S.; Pallarès, I.; Ventura, S. pH-Dependent Aggregation in Intrinsically Disordered Proteins Is Determined by Charge and Lipophilicity. Cells 2020, 9 (1), 145. 10.3390/cells9010145.

(46) Foster, P. H.; Ferry-Bolder, E.; Peakman, G.; Bouzigues, A.; Greaves, C. V.; Convery, R. S.; van Swieten, J. C.; Jiskoot, L. C.; Seelaar, H.; Moreno, F.; Sanchez-Valle, R.; Laforce Jr., R.; Graff, C.; Masellis, M.; Tartaglia, C.; Rowe, J. B.; Borroni, B.; Finger, E.; Synofzik, M.; Galimberti, D.; Vandenberghe, R.; Mendonca, A.; Butler, C.; Gerhard, A.; Ducharme, S.; Ber, I. L.; Tiraboschi, P.; Santana, I.; Pasquier, F.; Levin, J.; Otto, M.; Sorbi, S.; Rohrer, J. D.; Russell, L. L. Examining Longitudinal Changes of Disease Severity Scores in Familial Forms of Frontotemporal Dementia within the GENFI Cohort. Alzheimer’s & Dementia 2023, 19 (S18), e076588. 10.1002/alz.076588.

(47) Miyagawa, T.; Brushaber, D.; Syrjanen, J.; Kremers, W.; Fields, J.; Forsberg, L. K.; Heuer, H. W.; Knopman, D.; Kornak, J.; Boxer, A.; Rosen, H. J.; Boeve, B. F.; Appleby, B.; Bordelon, Y.; Bove, J.; Brannelly, P.; Caso, C.; Coppola, G.; Dever, R.; Dheel, C.; Dickerson, B.; Dickinson, S.; Dominguez, S.; Domoto-Reilly, K.; Faber, K.; Ferrell, J.; Fishman, A.; Fong, J.; Foroud, T.; Gavrilova, R.; Gearhart, D.; Ghazanfari, B.; Ghoshal, N.; Goldman, J. S.; Graff-Radford, J.; Graff-Radford, N.; Grant, I.; Grossman, M.; Haley, D.; Hsiung, R.; Huey, E.; Irwin, D.; Jones, D.; Jones, L.; Kantarci, K.; Karydas, A.; Kaufer, D.; Kerwin, D.; Kraft, R.; Kramer, J.; Kukull, W.; Litvan, I.; Lucente, D.; Lungu, C.; Mackenzie, I.; Maldonado, M.; Manoochehri, M.; McGinnis, S.; McKinley, E.; Mendez, M. F.; Miller, B.; Multani, N.; Onyike, C.; Padmanabhan, J.; Pantelyat, A.; Pearlman, R.; Petrucelli, L.; Potter, M.; Rademakers, R.; Ramos, E. M.; Rankin, K.; Rascovsky, K.; Roberson, E. D.; Rogalski, E.; Sengdy, P.; Shaw, L.; Tartaglia, M. C.; Tatton, N.; Taylor, J.; Toga, A.; Trojanowski, J. Q.; Wang, P.; Weintraub, S.; Wong, B.; Wszolek, Z. Utility of the Global CDR® plus NACC FTLD Rating and Development of Scoring Rules: Data from the ARTFL/LEFFTDS Consortium. Alzheimer’s & Dementia 2020, 16 (1), 106–117. 10.1002/alz.12033.

(48) Staffaroni, A. M.; Clark, A. L.; Taylor, J. C.; Heuer, H. W.; Sanderson-Cimino, M.; Wise, A. B.; Dhanam, S.; Cobigo, Y.; Wolf, A.; Manoochehri, M.; Forsberg, L.; Mester, C.; Rankin, K. P.; Appleby, B. S.; Bayram, E.; Bozoki, A.; Clark, D.; Darby, R. R.; Domoto-Reilly, K.; Fields, J. A.; Galasko, D.; Geschwind, D.; Ghoshal, N.; Graff-Radford, N.; Grossman, M.; Hsiung, G.-Y.; Huey, E. D.; Jones, D. T.; Lapid, M. I.; Litvan, I.; Masdeu, J. C.; Massimo, L.; Mendez, M. F.; Miyagawa, T.; Pascual, B.; Pressman, P.; Ramanan, V. K.; Ramos, E. M.; Rascovsky, K.; Roberson, E. D.; Tartaglia, M. C.; Wong, B.; Miller, B. L.; Kornak, J.; Kremers, W.; Hassenstab, J.; Kramer, J. H.; Boeve, B. F.; Rosen, H. J.; Boxer, A. L.; ALLFTD Consortium. Reliability and Validity of Smartphone Cognitive Testing for Frontotemporal Lobar Degeneration. JAMA Netw Open 2024, 7 (4), e244266. 10.1001/jamanetworkopen.2024.4266.

(49) van Blitterswijk, M.; DeJesus-Hernandez, M.; Rademakers, R. How Do C9ORF72 Repeat Expansions Cause ALS and FTD: Can We Learn from Other Non-Coding Repeat Expansion Disorders? Curr Opin Neurol 2012, 25 (6), 689–700. 10.1097/WCO.0b013e32835a3efb.

(50) Gaweda-Walerych, K.; Aragona, V.; Lodato, S.; Sitek, E. J.; Narożańska, E.; Buratti, E. Progranulin Deficiency in the Brain: The Interplay between Neuronal and Non-Neuronal Cells. Translational Neurodegeneration 2025, 14 (1), 18. 10.1186/s40035-025-00475-8.

(51) Hodgson, R. E.; Rayment, J. A.; Huang, W.-P.; Avila, A. S.; Ellis, B. C. S.; Lin, Y.-H.; Soni, N.; Hautbergue, G. M.; Shelkovnikova, T. A. C9orf72 Poly-PR Forms Anisotropic Condensates Causative of Nuclear TDP-43 Pathology. iScience 2024, 27 (10). 10.1016/j.isci.2024.110937.

(52) Montembeault, M.; Brambati, S. M.; Gorno-Tempini, M. L.; Migliaccio, R. Clinical, Anatomical, and Pathological Features in the Three Variants of Primary Progressive Aphasia: A Review. Front. Neurol. 2018, 9. 10.3389/fneur.2018.00692.

