## Supplemental Information for "Digital seed amplification assay for TDP-43 aggregate quantification in CSF"

**Table of contents:**

Figure S1- Homoscedasticity plots of multiple linear regression performance with different weighing methods.

Figure S2- Representative transmission electron microscopy images of pre-formed fibrils.

Figure S3- Summarized results from a set of experimental variation of reaction parameters.

Figure S4- Image processing pipeline.

Figure S5- TDPC aggregate growth and quantification in microwells.

Figure S6- Simple linear regression of seed concentration vs. FTLDCDR score.

Figure S7- Observed seed concentrations of samples grouped by primary clinical phenotype.

Figure S8- Performance assessment plots for MLR.

Table S1- Replicate measurements and their corresponding coefficient of variation percentages of samples.

Table S2 – VIF table of variables in multilinear regression.

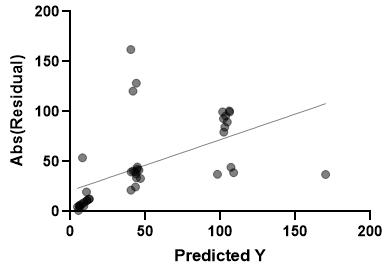

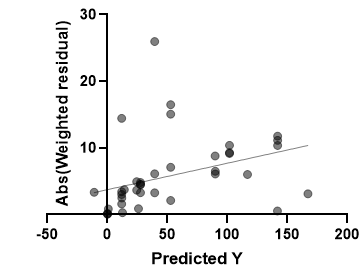

**A**

**B**

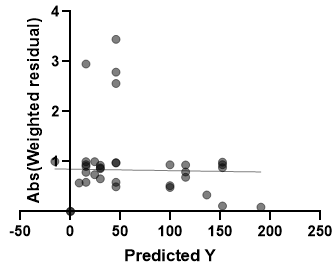

**C**

**Figure S1- Homoscedasticity plots of multiple linear regression performance with different weighing methods. A.** Unweighted**. B**. Weight by 1/Y**. C.** Weight by 1/Y^2^**.**

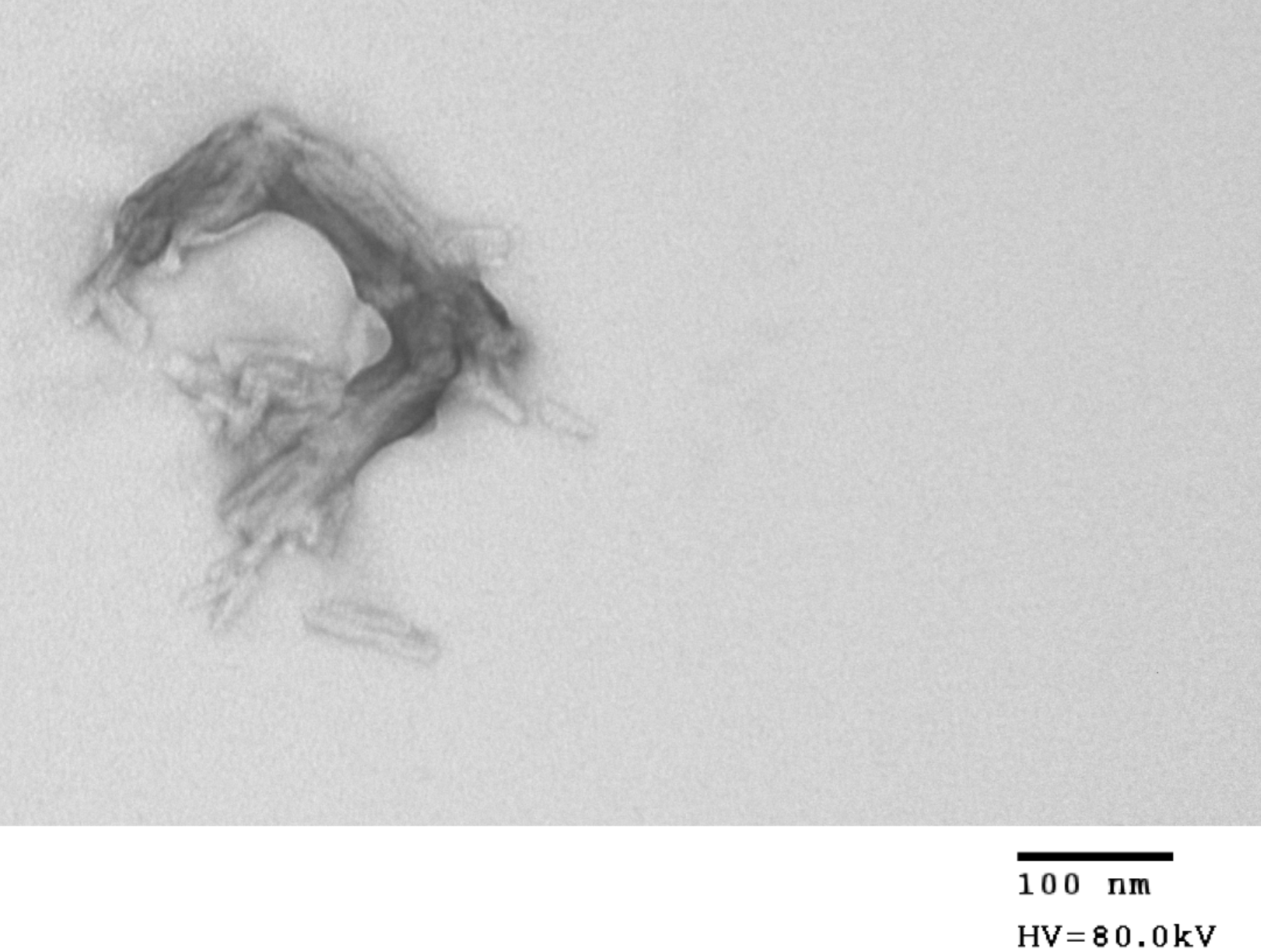

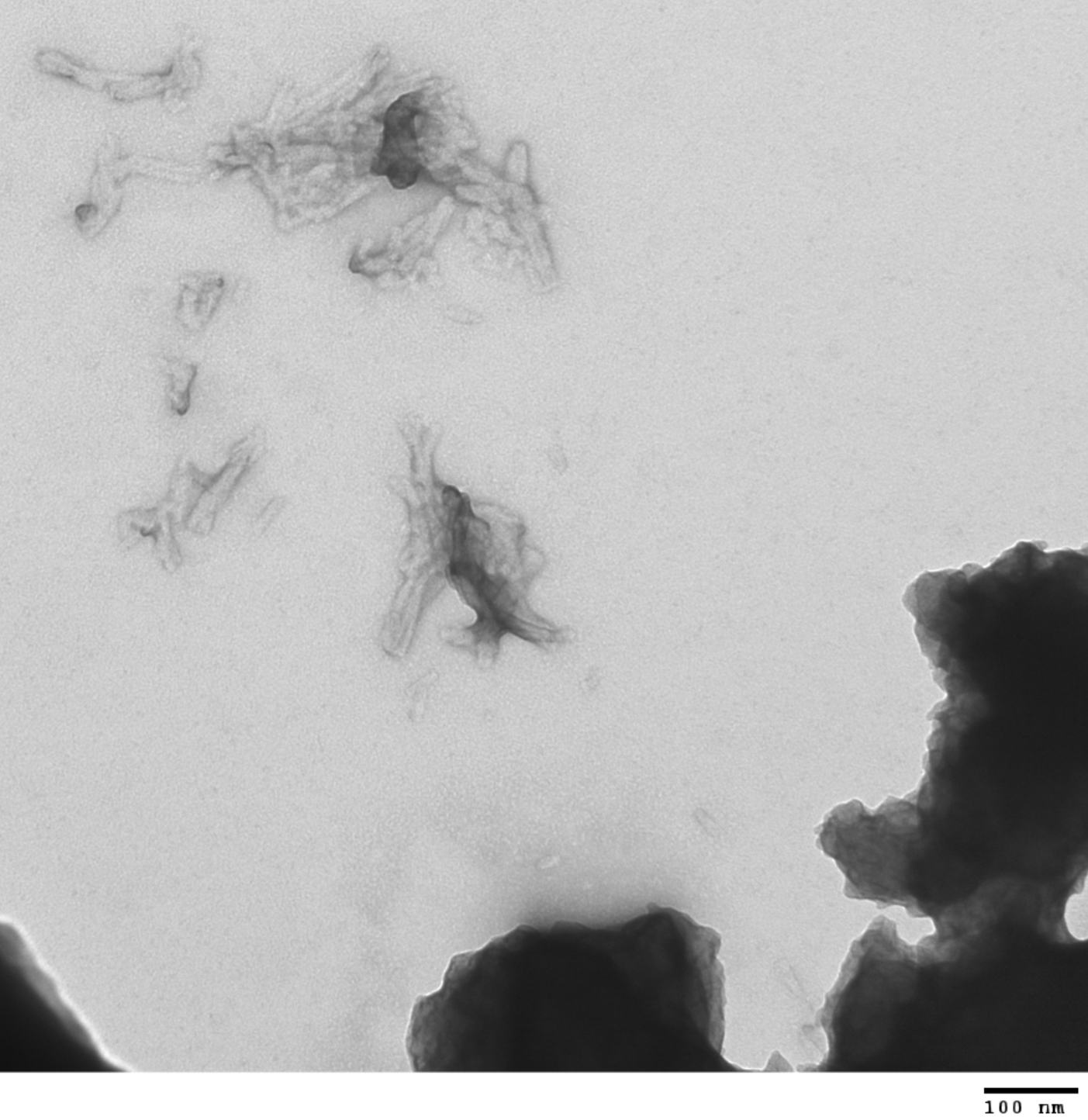

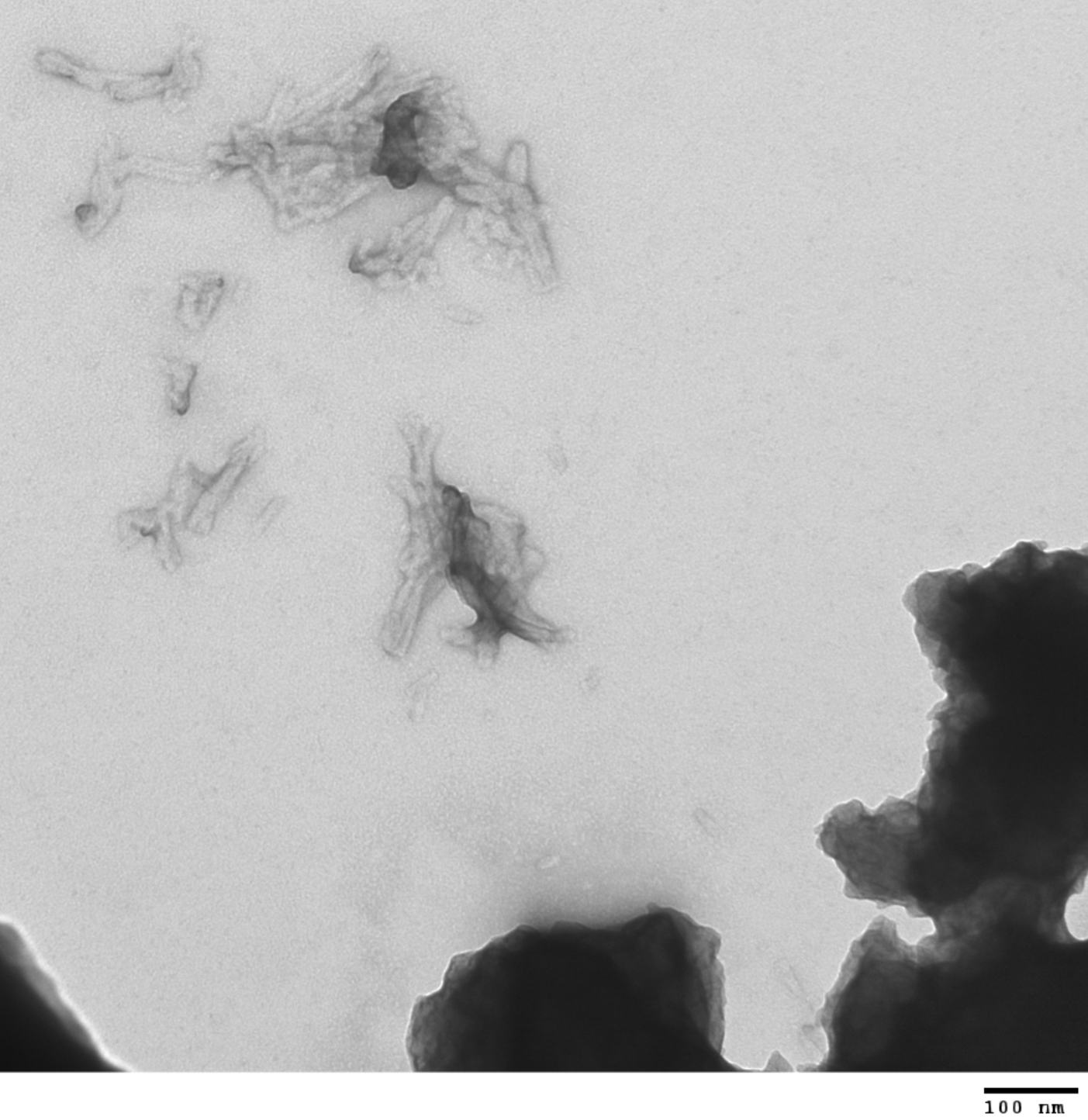

**Figure S2- Representative transmission electron microscopy images of pre-formed fibrils (PFFs).** PFFs were generated from TDP-C to serve as dSAA reaction seeds. Scale bar: 100 nm.

| 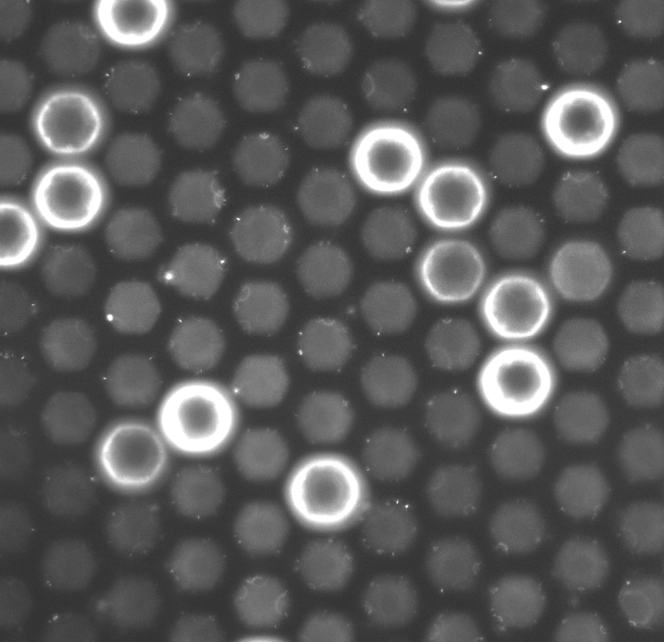 | 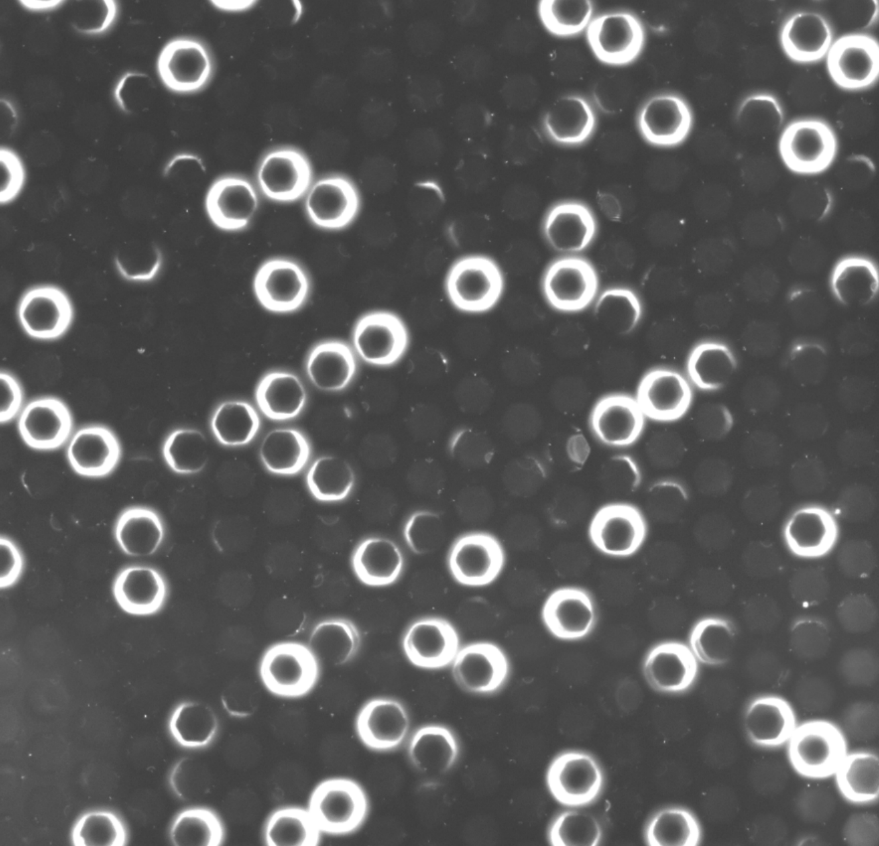 | 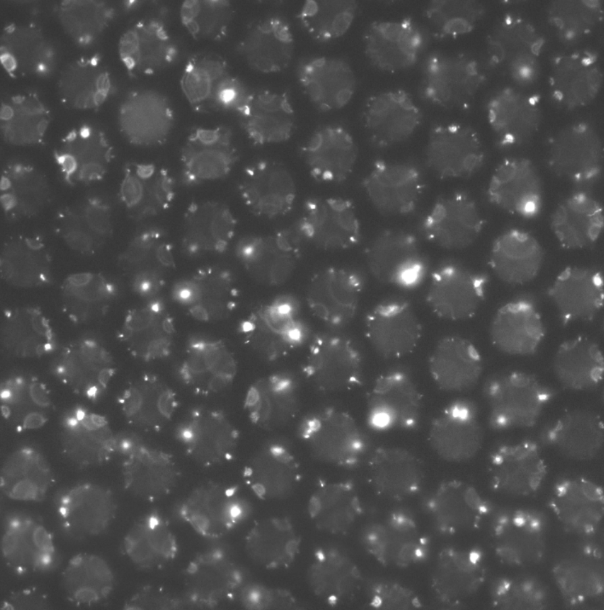 | 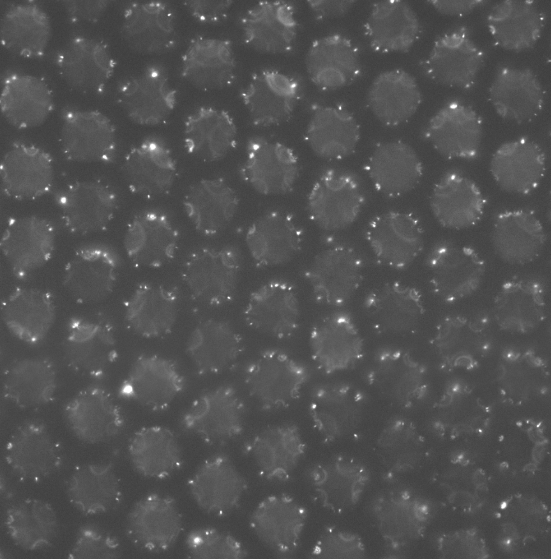 |
| --- | --- | --- | --- |
| SNR 0.14  PBS 0.1M pH 7.5, GdnCl 0.1M, 0.1% BSA, 0.02% triton, 0.0015% SDS, 2uM X34, 10 nM PFFs | SNR 0.88  TRIS 0.1M pH 8, GdnCl 0.1M, 0.1% BSA, 0.02% triton, 0.0015% SDS, 2uM X34, 10 nM PFFs | SNR 0.97  PIPES 0.1M pH 7.5, GdnCl 0.1M, 0.1% BSA, 0.02% triton, 0.0015% SDS, 2uM X34, 10 nM PFFs | SNR 0.96  PBS 0.1M pH 7.5, NaCl 0.1M, 0.1% BSA, 0.02% triton, 0.0015% SDS, 2uM X34, 10 nM PFFs |
| 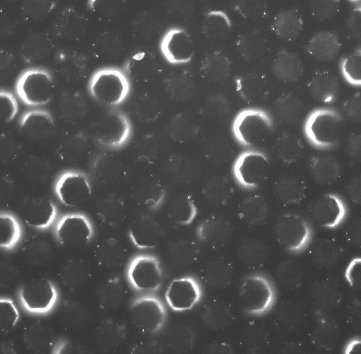 | 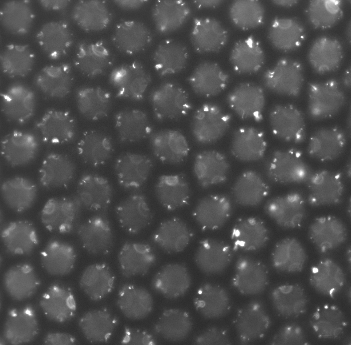 | 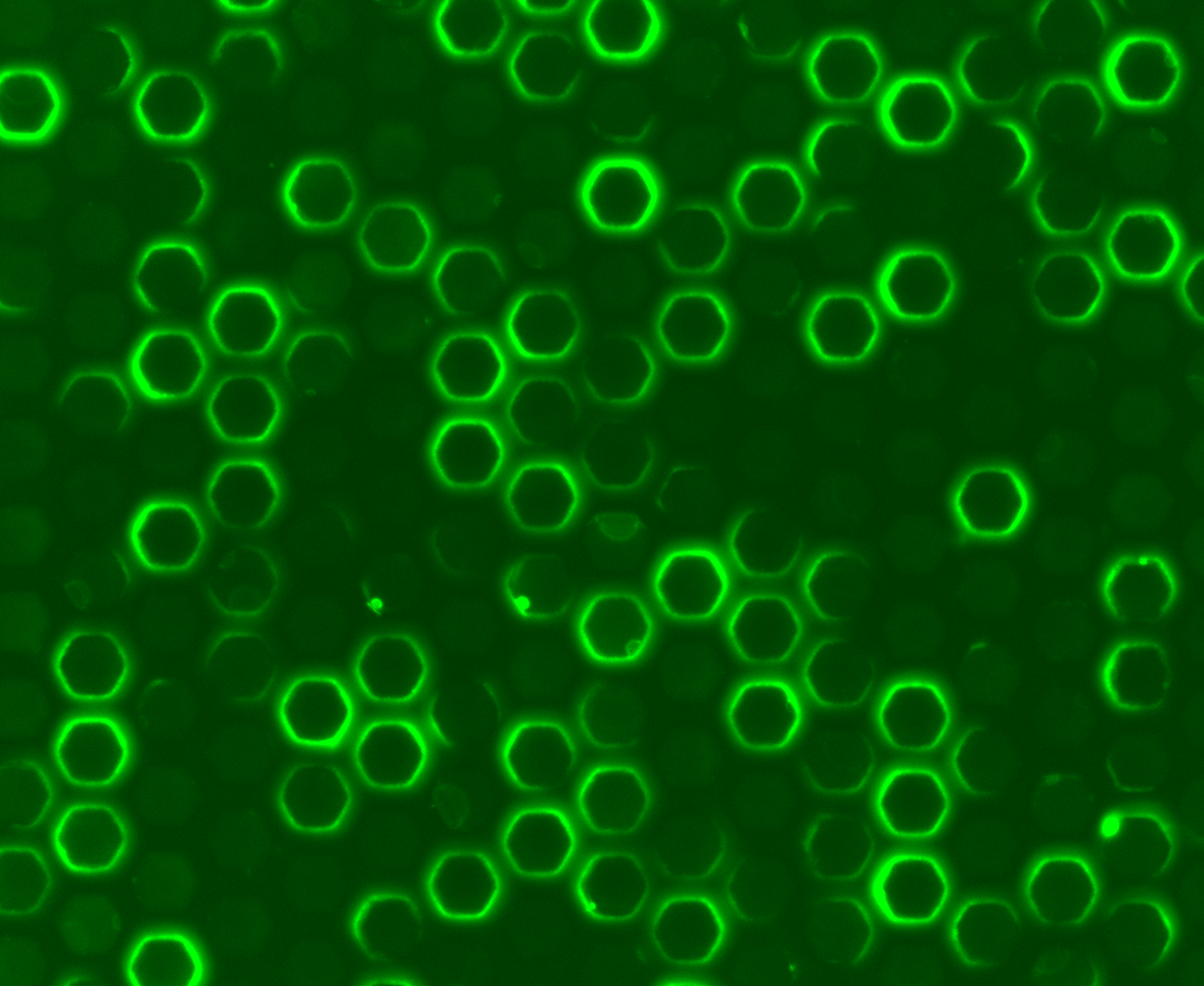 | 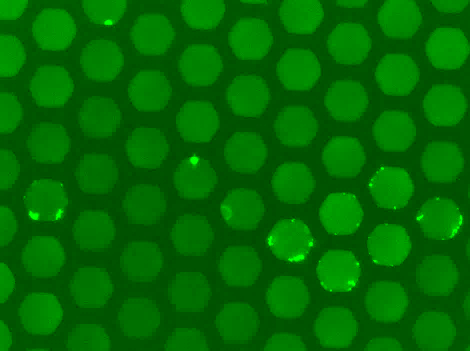 |
| SNR 1.21  TRIS 0.1M pH 8, GdnCl 0.1M, 0.1% BSA, 0.02% triton, 0.0015% SDS, 2uM X34, 10 nM PFFs | SNR 1.07  PIPES 0.1M pH 7.5, GdnCl 0.1M, 0.1% BSA, 0.02% triton, 0.0015% SDS, 2uM X34, 10 nM PFFs | SNR 1.01  TRIS 20mM pH 7.5, GdnCl 0.1M, 0.1% BSA, 0.02% triton, 0.0015% SDS, 2uM X34, 10 nM PFFs | SNR 3.60  PIPES 100mM pH 7.5, GdnCl 100mM, 0.1% BSA, 0.02% triton, 0.0015% SDS, 5mM DTT, 2uM X34, 10 nM PFFs |
| 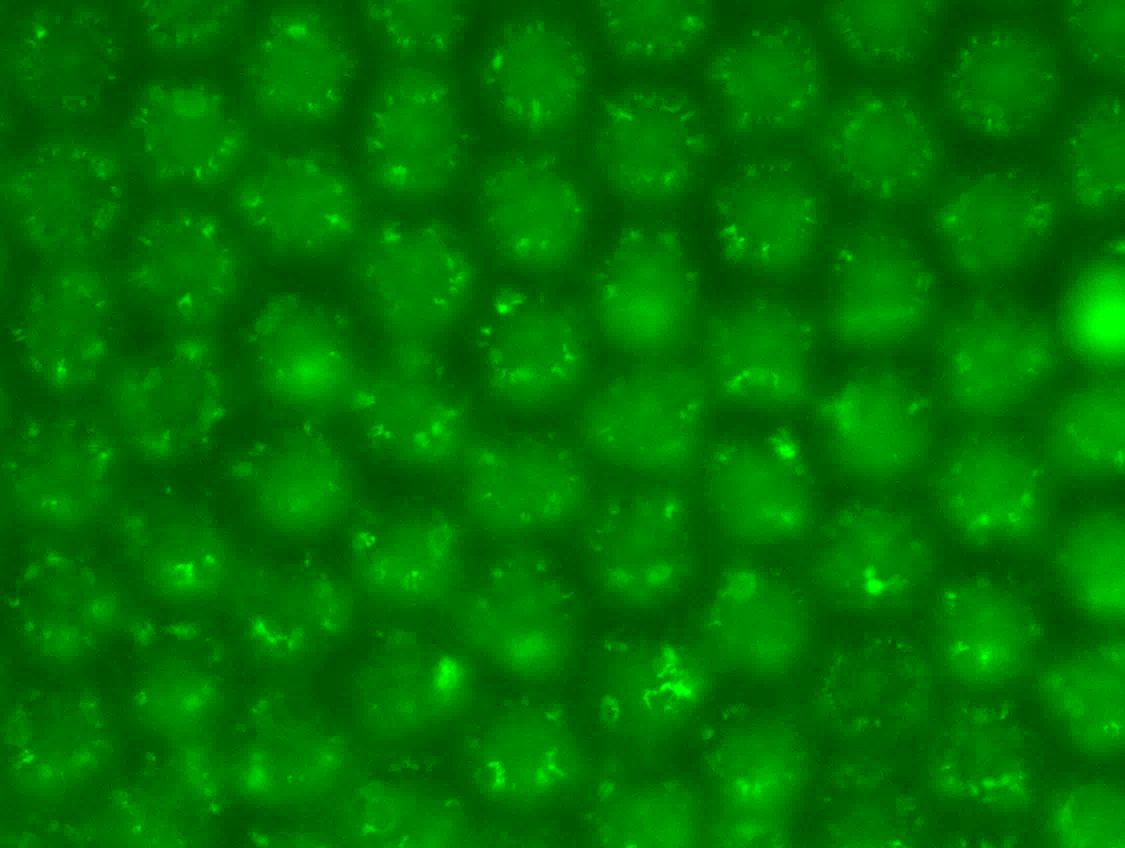 | 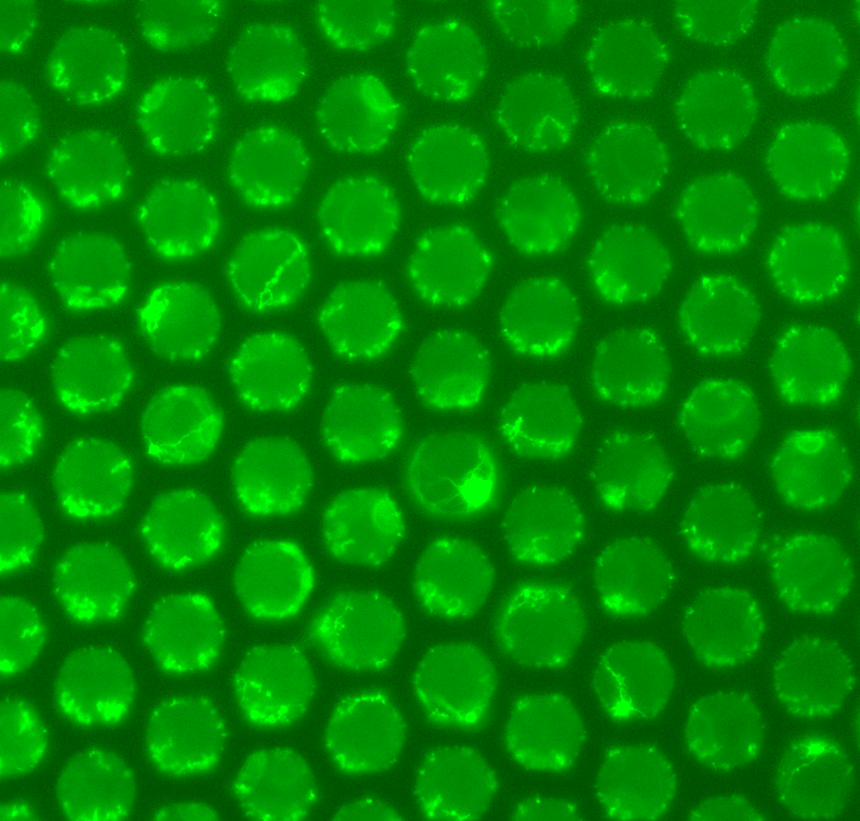 | 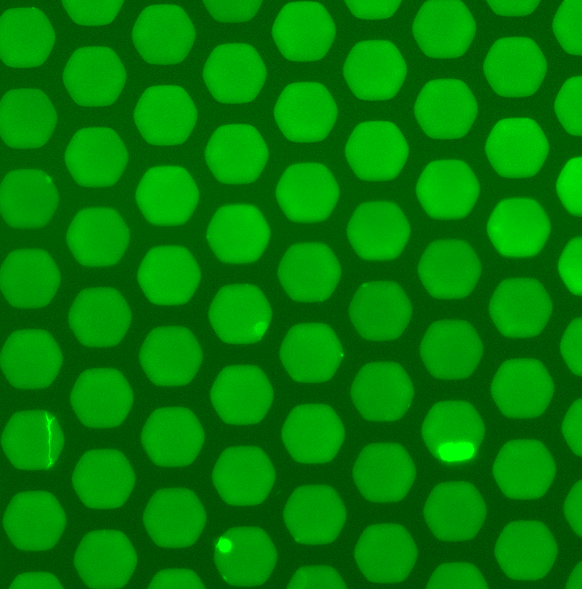 | 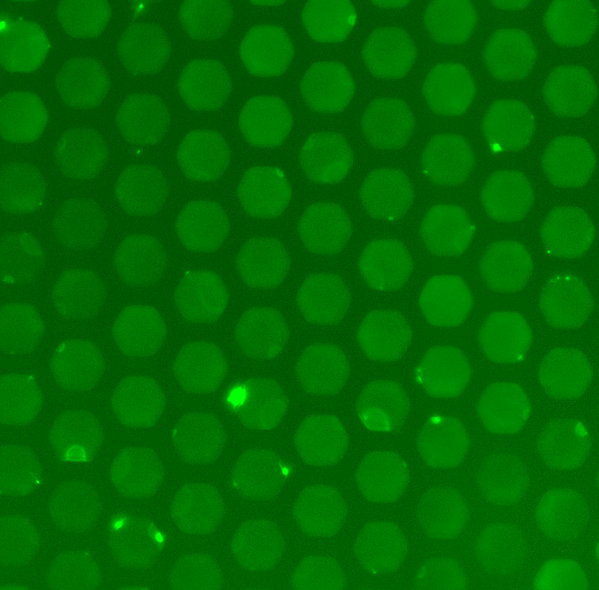 |
| SNR 15.00  TRIS 20mM pH 7.5, GdnCl 100mM, 0.1% BSA, 0.02% triton, 0.0015% SDS, 5mM DTT, 2uM X34, 10 nM PFFs | SNR 9.09  TRIS 20mM pH 7.5, GdnCl 100mM, 0.1% BSA, 0.02% triton, 0.00015% SDS, 10mM DTT, 2uM X34, 10 nM PFFs | SNR 2.50  TRIS 20mM pH 6.5, GdnCl 100mM, 0.1% BSA, 0.02% triton, 0.00015% SDS, 10mM DTT, 2uM X34, 10 nM PFFs | SNR 1.43  TRIS 20mM pH 7.5, GdnCl 100mM, 0.1% BSA, 0.1% triton, 0.00015% SDS, 10mM DTT, 2uM X34, 10 nM PFFs |
| 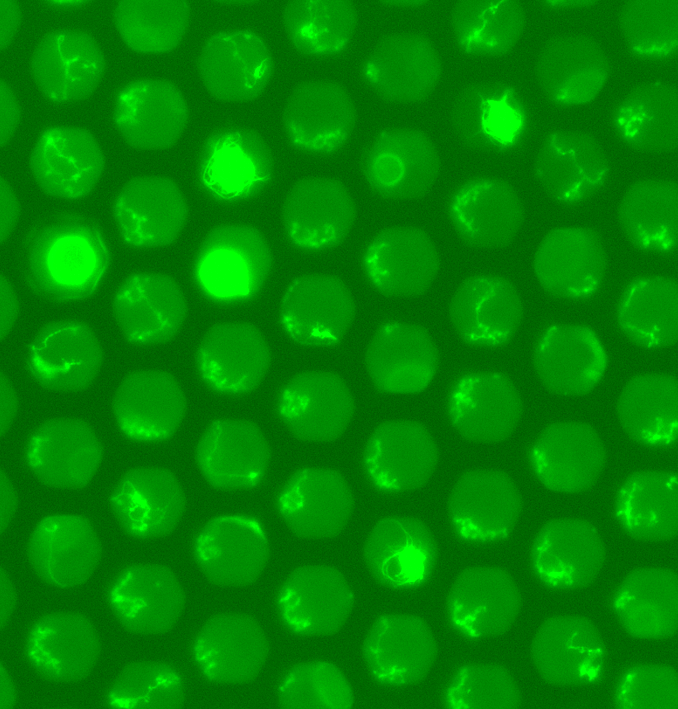 | 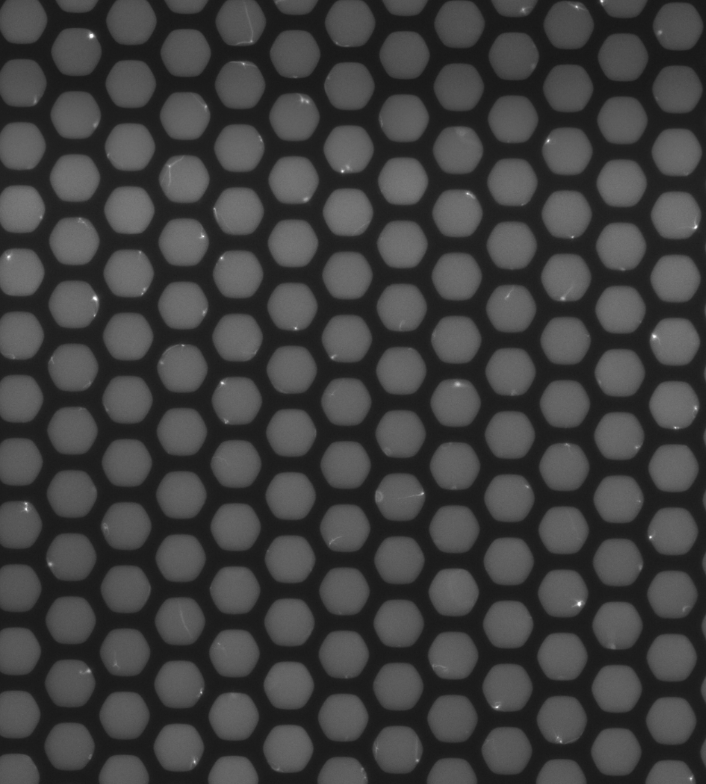 | 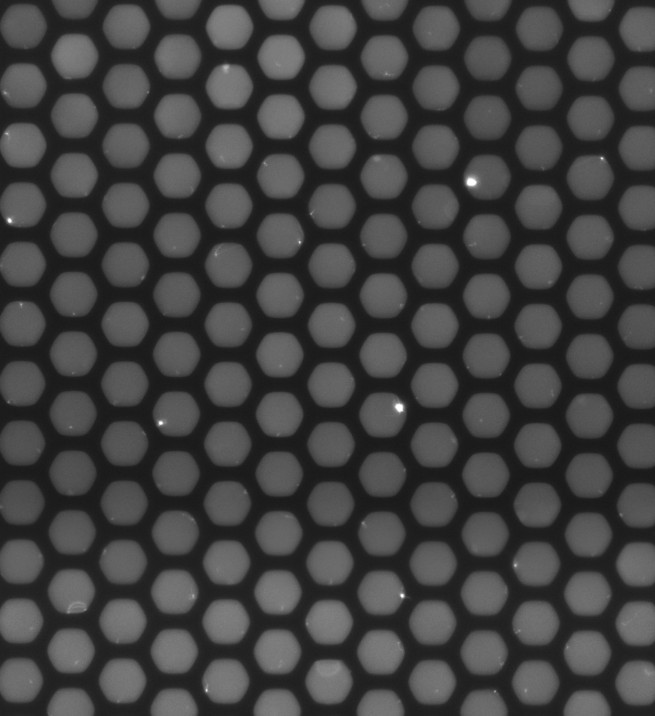 | 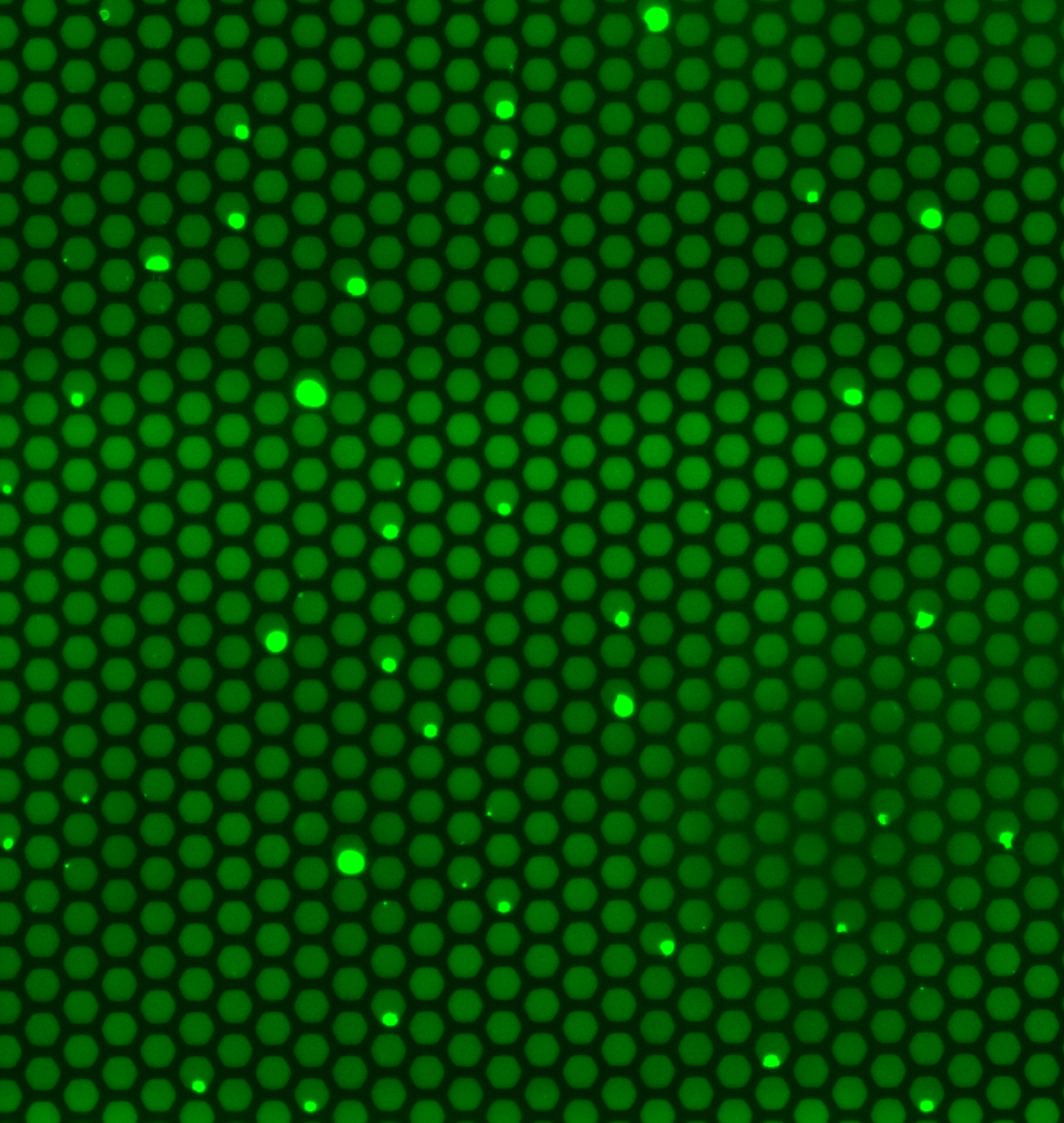 |
| SNR 24.00  TRIS 20mM pH 6.5, GdnCl 0.1M, 0.1% BSA, 0.1% triton, 0.00015% SDS, 10mM DTT, 2uM X34, 10 nM PFFs | SNR 0.56  TRIS 20mM pH 6.5, GdnCl 0.1M, 0.1% triton, no BSA, 10mM DTT, 5uM X34, 10 nM PFFs | SNR 1.95  TRIS 20mM pH 6.5, NaCl 0.1M, 0.1% triton, no BSA, 5mM DTT, 0.0015% SDS, 5uM X34, 10 nM PFFs | SNR 8.01  TRIS 20mM pH 6.5, GdnCl 0.1M, 0.1% triton, no BSA, 10mM DTT, 0.0015% SDS, 5uM X34, 10 nM PFFs |

**Figure S3- Summarized SNR of a set of experimental variations of reaction parameters.** Representative images of microwell arrays at t = 15 hours, reaction conditions below each image. Scale bar: 60 μm. SNR is calculated as on-well% of reaction seeded with 10 nM PFFs divided by the on-well% of the unseeded reaction with the same conditions. Abbreviations: signal to noise ratio; SNR, phosphate-buffered saline; PBS, Tris(hydroxymethyl)aminomethane; TRIS, 1,4-piperazinediethanesulfonic acid; PIPES, sodium dodecyl sulfate; SDS, guanidine hydrochloride; GdnCl, bovine serum albumin; BSA, dithiothreitol; DTT, pre-formed fibrils; PFFs.

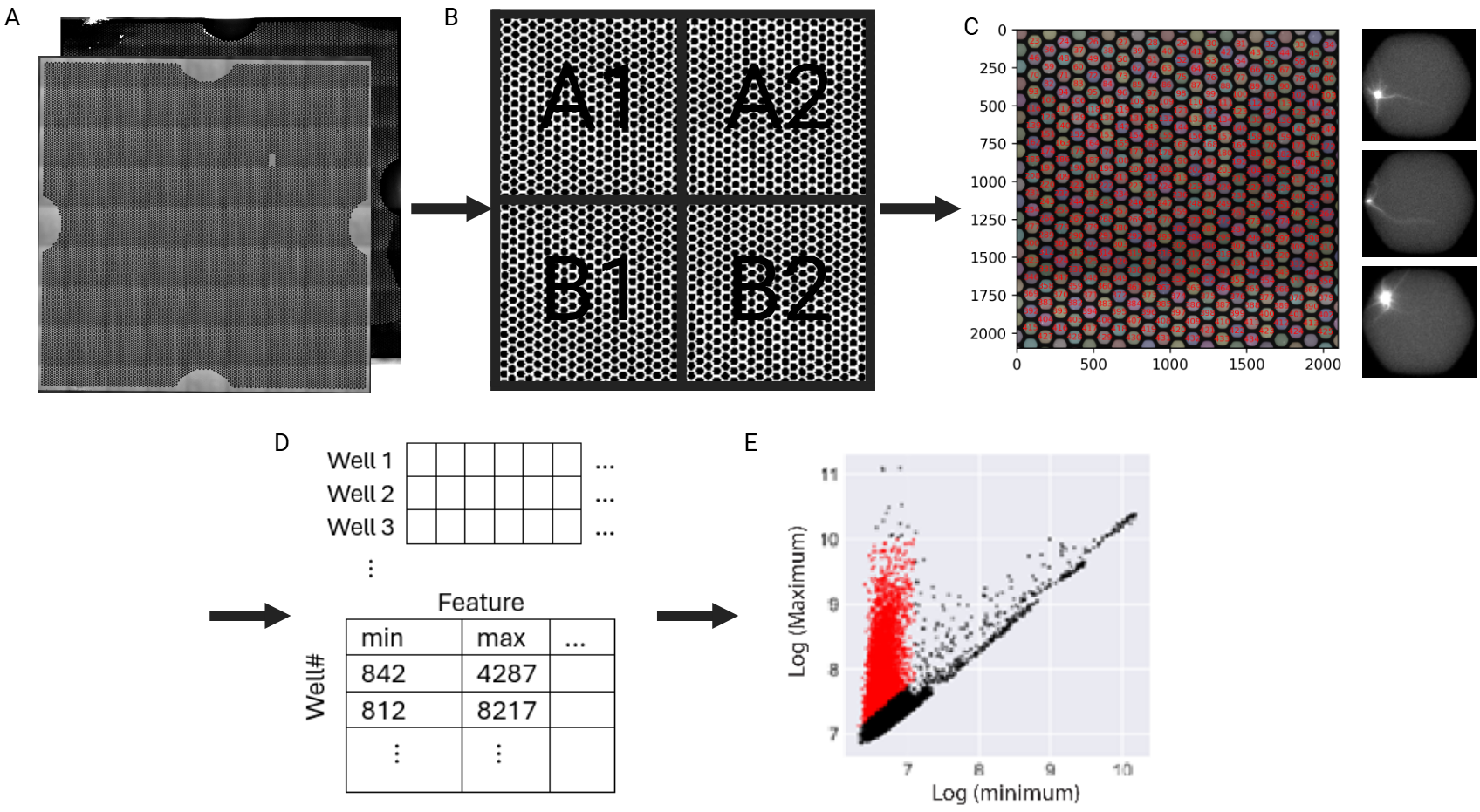

**Figure S4- Image processing pipeline.** On-wells percentages in each chip were determined using a custom Python image processing pipeline analyzing approximately 13,500 wells. **A.** Raw images include two channels, a fluorescence channel and a brightfield channel. **B.** Brightfield channel was binarized, sectioned. **C.** and used to detect the center point of each well within sections **(left)**. Then, each well was individually extracted from the fluorescence channel as a separate 90x90 pixel image **(right)**. **D.** Each image was reshaped to be a 1-dimensional vector with 8100 values. All images were then combined into a single matrix W of dimension (n_wells, 8100) **(top)**, and features were generated (such as well minimum, maximum, etc.) in a separate metadata matrix M with dimension (n_wells, n_features) **(bottom)**. **E.** Features were then used for initial well classification to create a model training set for a generalized classification model **(E)**.

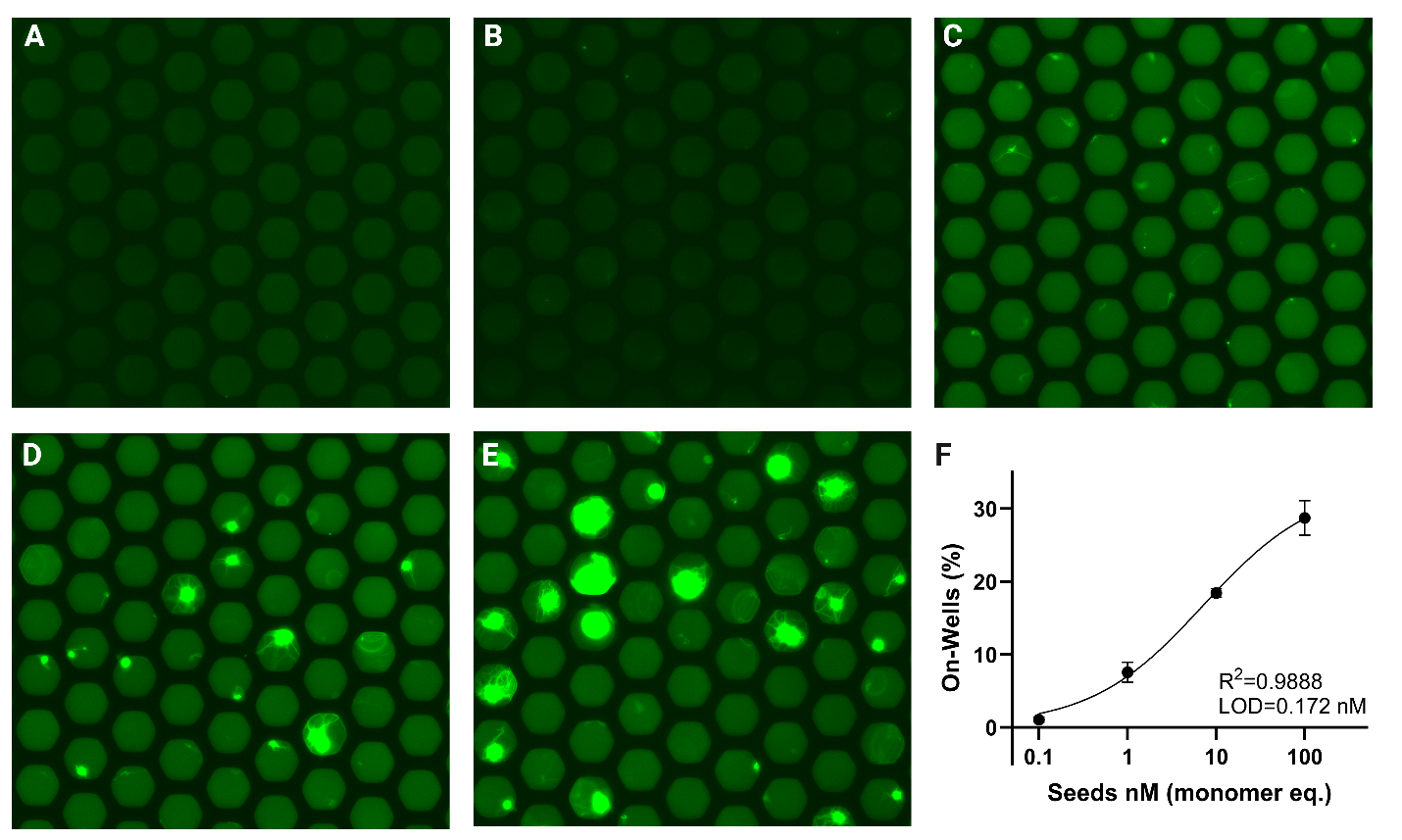

**Figure S5- TDPC aggregate growth and quantification in microwells.** Representative images of microwell arrays at t = 15 hours in healthy brain lysate **(A-E)**. The SAA reaction for PFFs used as reaction seeds concentrations of 0 nM **(A)**, 0.1nM **(B)** 1 nM **(C)**, 10 nM **(D)**, and 100 nM **(E)**. The percentage of wells with growing aggregates vs. the concentrations of pre-formed filaments used as reaction seeds spiked into healthy brain lysate, trend line shows the 4PL curve **(F)**.

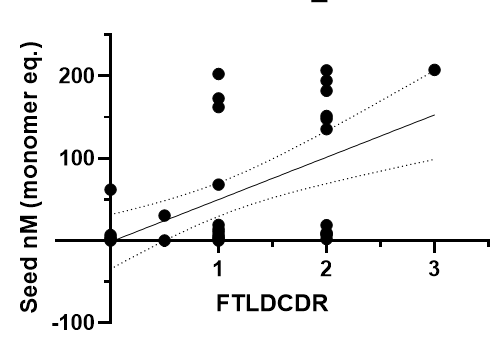

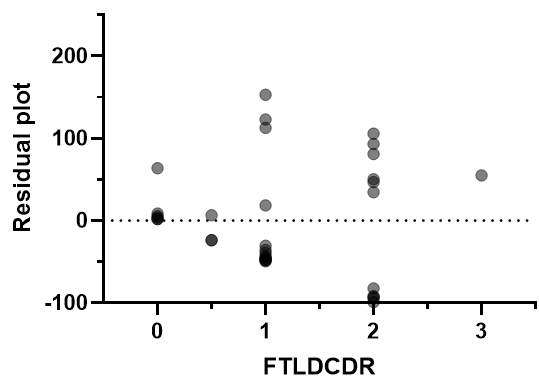

**Figure S6-** **Simple linear regression of seed concentration vs. FTLDCDR score. A.** Seed concentration vs. FTLDCDR score plot. **B.** Residual plot.

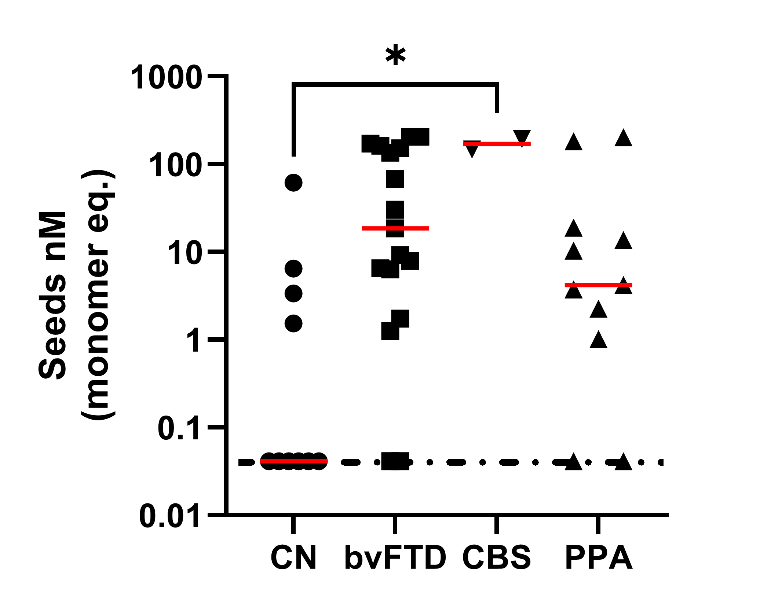

**Figure S7- Observed seed concentrations of samples grouped by primary clinical phenotype.** The primary clinical phenotype was determined through the clinician’s diagnostic impression, assessed through the National Alzheimer's Coordinating Center B9 form known as Clinician’s Judgement of Symptoms. Red horizontal lines represent group median value. Dotted horizontal line represents assay LOD. To assess the significance of median variance between binned categories vs. control one-way ANOVA with post-doc Tuckey’s test was used.

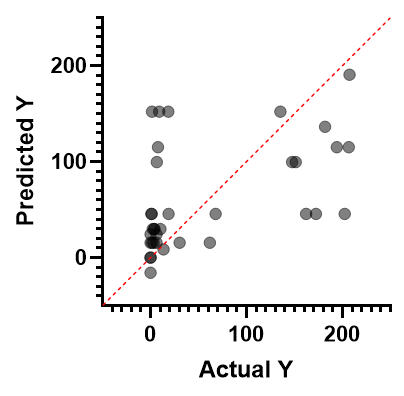

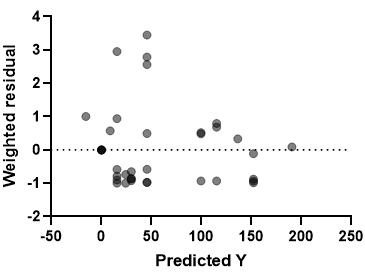

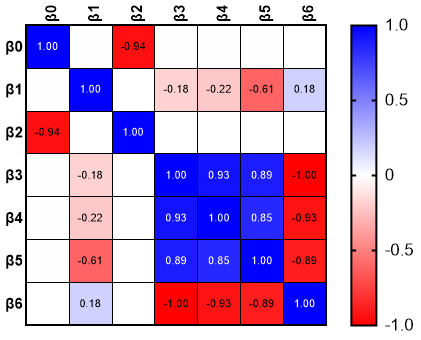

**C**

**B**

**A**

**Figure S8- Performance assessment plots for MLR. A.** Actual vs. predicted plot. **B.** Residual plot**. C.** Parameter covariance table. β0- Intercept, β1- Sex[F], β2- Age at CSF collection, β3- FTLD Subtype[C9ORF72], β4- FTLD Subtype[GRN], β5- FTLD Subtype[NONE-PPA].

**Table S1- Replicate measurements and their corresponding coefficient of variation percentages of samples.**

|  | **On-well %** | | | | | |
| --- | --- | --- | --- | --- | --- | --- |
|  | **Replicate 1** | **Replicate 2** | **Replicate 3** | **Mean** | **SD** | **CV** |
| **S01** | 3.81 | 0.58 | 1.34 | 1.91 | 1.38 | 0.72 |
| **S02** | 1.65 | 0.28 | 0.35 | 0.76 | 0.63 | 0.83 |
| **S03** | 1.28 | 3.95 | 4.99 | 3.40 | 1.56 | 0.46 |
| **S04** | 0.16 | 2.57 | 16.68 | 6.47 | 7.29 | 1.13 |
| **S05** | 3.39 | 0.21 | 0.32 | 1.30 | 1.47 | 1.13 |
| **S06** | 0.53 | 0.24 | 0.69 | 0.49 | 0.19 | 0.39 |
| **S07** | 1.28 | 1.02 | 0.34 | 0.88 | 0.40 | 0.45 |
| **S08** | 0.73 | 10.35 | 12.67 | 7.92 | 5.17 | 0.65 |
| **S09** | 1.65 | 0.63 | 2.19 | 1.49 | 0.65 | 0.43 |
| **S10** | 97.56 | 90.52 | 90.77 | 92.95 | 3.26 | 0.04 |
| **S11** | 14.44 | 67.98 | 77.68 | 53.37 | 27.81 | 0.52 |
| **S12** | 72.39 | 90.57 | 56.22 | 73.06 | 14.03 | 0.19 |
| **S13** | 89.55 | 20.14 | 75.87 | 61.85 | 30.01 | 0.48 |
| **S14** | 87.33 | 88.79 | 97.07 | 91.06 | 4.29 | 0.05 |
| **S15** | 67.59 | 91.74 | 86.25 | 81.86 | 10.34 | 0.13 |
| **S16** | 86.25 | 85.59 | 90.27 | 87.37 | 2.07 | 0.02 |
| **S17** | 40.07 | 67.79 | 96.92 | 68.26 | 23.21 | 0.34 |
| **S18** | 13.77 | 7.67 | 9.39 | 10.27 | 2.57 | 0.25 |
| **S19** | 0.60 | 0.16 | 0.19 | 0.32 | 0.20 | 0.64 |
| **S20** | 2.61 | 3.50 | 0.98 | 2.37 | 1.05 | 0.44 |
| **S21** | 1.04 | 0.19 | 0.33 | 0.52 | 0.38 | 0.72 |
| **S22** | 1.17 | 0.36 | 1.16 | 0.90 | 0.38 | 0.42 |
| **S23** | 1.00 | 10.13 | 0.23 | 3.79 | 4.50 | 1.19 |
| **S24** | 0.35 | 1.19 | 0.52 | 0.69 | 0.36 | 0.53 |
| **S25** | 0.70 | 0.39 | 0.88 | 0.66 | 0.21 | 0.31 |
| **S26** | 1.00 | 0.76 | 0.99 | 0.92 | 0.11 | 0.12 |
| **S27** | 0.63 | 1.64 | 2.92 | 1.73 | 0.94 | 0.54 |
| **S28** | 43.50 | 77.11 | 5.67 | 42.09 | 29.18 | 0.69 |
| **S29** | 68.73 | 71.04 | 93.34 | 77.70 | 11.10 | 0.14 |
| **S30** | 52.31 | 52.65 | 94.30 | 66.42 | 19.72 | 0.30 |
| **S31** | 2.69 | 15.90 | 34.23 | 17.61 | 12.93 | 0.73 |
| **S32** | 4.07 | 2.06 | 6.53 | 4.22 | 1.83 | 0.43 |
| **S33** | 6.53 | 12.53 | 0.50 | 6.52 | 4.91 | 0.75 |
| **S34** | 0.84 | 6.47 | 11.64 | 6.32 | 4.41 | 0.70 |
| **S35** | 89.72 | 93.26 | 96.79 | 93.25 | 2.88 | 0.03 |
| **S36** | 21.67 | 15.24 | 3.19 | 13.37 | 7.66 | 0.57 |
| **S37** | 8.07 | 21.12 | 49.30 | 26.16 | 17.21 | 0.66 |
| **S38** | 21.14 | 3.06 | 3.32 | 9.17 | 8.46 | 0.92 |
| **S39** | 1.00 | 27.85 | 24.93 | 17.93 | 12.03 | 0.67 |
| **S40** | 0.68 | 0.90 | 1.00 | 0.86 | 0.13 | 0.15 |

**Table S2 – VIF table of variables in multilinear regression**

| **Variable** | | **VIF** | | **R^2^ with other variables** |
| --- | --- | --- | --- | --- |
| **Female** | | 1.049 | | 0.046 |
| **Age at CSF collection** | | 1.239 | | 0.193 |
| **FTLD Subtype** | **C9ORF72** | 3.331 | 0.700 | |
|  | **GRN** | 4.981 | 0.799 | |
|  | **Sporadic-PPA** | 2.685 | 0.628 | |
| **FTLDCDR** | | 3.144 | | 0.682 |
