## Supplementary material for "Digital seed amplification assay for TDP-43 aggregate quantification in CSF": Table 1

| **Characteristic** | ***C9ORF72*** | ***GRN*** | **Sporadic PPA** | **Clinically Normal** | **Totals patients** |
| --- | --- | --- | --- | --- | --- |
| **Female, *n*. (%)** | 5 (50) | 5 (50) | 5 (50) | 5 (50) | 20 (50) |
| **Age at CSF collection, y. (±SD)** | 58 (±10) | 61 (±7) | 59 (±10) | 50 (±14) | 57 (±10) |
| **Age at onset, y. (±SD)** | 50 (±11) | 58 (±6) | 50 (±13) | - | 53 (±11) * |
| **Onset-CSF collection, y. (±SD)** | 9 (±6) | 3 (±2) | 8 (±10) | - | 4 (±8) * |
| **Primary clinical phenotype** |  |  |  |  |  |
| **bvFTD, n.** | 10 | 7 | 0 | 0 | 17 |
| **PPA, *n*.** | 0 | 1 | 10 | 0 | 11 |
| **CBS, *n*.** | 0 | 2 | 0 | 0 | 2 |
| **Secondary clinical phenotype** |  |  |  |  |  |
| **MCI, n.** | 2 | 0 | 0 | 0 | 2 |
| **AD, n.** | 0 | 1 | 0 | 0 | 1 |
| **CBS, n.** | 0 | 1 | 0 | 0 | 1 |
| **Total, *n*.** | 10 | 10 | 10 | 10 | 40 |
| **Positive dSAA, *n*. (%)** | 9 (90) | 9 (90) | 8 (80) | 4 (40) | 30 (75) |

* Values exclude CN population
