## Supplementary material for "Digital seed amplification assay for TDP-43 aggregate quantification in CSF": Table 3

| **Degrees of Freedom** | **Multiple R** | **R^2^ (weighted)** | | **Adjusted R^2^**  **(weighted)** | | **Weighted Sum of Squares**  **(1/Y²)** | | | **Sy.x** | **RMSE** | | **AICc** |
| --- | --- | --- | --- | --- | --- | --- | --- | --- | --- | --- | --- | --- |
| 33 | 0.617 | 0.380 | | 0.268 | | 53.7 | | | 1.28 | 1.17 | | 32.4 |
|  | | | **SS** | | **DF** | | **MS** | **F-statistic** | | | ***p*-Value** | |
| **Regression** | | | 32.9 | | 6 | | 5.49 | 3.37 | | | 0.0105 | |
| **Female** | | | 8.95 | | 1 | | 8.95 | 5.50 | | | 0.0252 | |
| **Age at CSF collection, y.** | | | 1.51x10^-7^ | | 1 | | 1.51x10^-7^ | 9.28x10^-8^ | | | 0.100 | |
| **FTLD Subtype** | | | 32.9 | | 3 | | 11.0 | 6.75 | | | 0.0011 | |
| **FTLDCDR** | | | 25.5 | | 1 | | 25.5 | 15.7 | | | 0.0004 | |
| **Residual** | | | 53.7 | | 33 | | 1.63 |  | | |  | |
| **Total** | | | 86.6 | | 39 | |  |  | | |  | |
