## Supplementary material for "Digital seed amplification assay for TDP-43 aggregate quantification in CSF": Table 4

| **Predictor** | | **Estimate** | **\|t\|** | ***p*-Value** | **95% CI Lower** | **95% CI Upper** |
| --- | --- | --- | --- | --- | --- | --- |
| **Intercept** | | 0.0410 | 0.582 | 0.565 | -0.102 | 0.184 |
| **Female** | | 17.5 | 2.34 | 0.0252 | 2.32 | 32.7 |
| **Age at CSF collection, y.** | | 4.69ex10^-7^ | 0.000305 | 1.00 | -0.00313 | 0.00313 |
| **FTLD Subtype** | **C9ORF72** | -44.8 | 3.96 | 0.0004 | -67.8 | -21.8 |
|  | **GRN** | -80.8 | 3.12 | 0.0037 | -133 | -28.2 |
|  | **Sporadic svPPA** | -62.3 | 4.18 | 0.0002 | -92.6 | -32.0 |
| **FTDCDR** | | 89.6 | 3.96 | 0.0004 | 43.5 | 136 |
